# Assessing the Impacts of COVID-19 and Social Isolation on Mental Health in the United States of America

**DOI:** 10.1101/2022.09.09.22277383

**Authors:** Alexander Fulk, Raul Saenz-Escarcega, Hiroko Kobayashi, Innocent Maposa, Folashade Agusto

## Abstract

The COVID-19 pandemic has had a devastating impact on the world at large with over 500 million cases and over 6 million deaths reported thus far. Of those, over 85 million cases and 1 million deaths have occurred in the United States of America. The mental health of the general population has been impacted by several aspects of the pandemic including lockdowns, media sensationalism, social isolation, and spread of the disease. In this paper, we examine the effect that social isolation and COVID-19 infection and related death had on the prevalence of anxiety and depression in the general population of the USA in a state-by-state multiple time-series analysis. Vector Error Correction Models are estimated and we subsequently evaluated the coefficients of the estimated models and calculated their impulse response functions for further interpretation. We found that variables related to COVID-19 overall led to increase in both anxiety and depression across the studied period, while variables related to social isolation had a varied effect depending on the state being considered.

## 1 Introduction

Since its emergence, the World Health Organization (WHO) has reported over 500 million cases of COVID-19 globally and over 6 million deaths [1]. COVID-19 is a novel coronavirus disease caused by the severe acute respiratory syndrome coronavirus 2 (SARS-Cov-2) that set off an unprecedented global health crisis creating a myriad of lockdown restrictions in response to the surge of COVID-19 cases in different parts of the world. First discovered in Wuhan, China in December 2019, the virus has spread from person to person due to its respiratory transmissibility. Individuals infected with the virus can pass it on to susceptible individuals by being within 2 meters of contact, sneezing, and/or coughing through aerosol droplets containing the virus. People could also become infected if they touch their eyes or nose after coming into contact with contaminated surfaces [2]. Areas with poor ventilation in indoor settings and/or crowded rooms can allow for spread to susceptible individuals due to aerosols of the virus suspended for longer in these specific conditions [4]. Reported symptoms of COVID-19 range from mild to severe symptoms including: fever, sore throat, diarrhea, shortness of breath, headache, and body aches, and there are reports of asymptomatic individuals who experience no symptoms, but can still pass on the virus. Older populations (*>*65+) or those with other underlying medical conditions are associated with a higher likelihood of severe symptoms from COVID-19 [5].

In response to the early outbreaks and surges of COVID-19 cases, several country leaders and government officials set up lockdown mandates to suppress transmission between people [6, 7]. These restrictions were in place to stop the ever-growing incidence of COVID-19 cases and death from rising drastically. However, the lockdown mandates led to other unprecedented effects on the mental health of the public [7]. With lockdown mandates becoming prominent worldwide, there was a rise in mental health problems from pre-pandemic data [8] and an increase in general worry related to the pandemic [9]. Culminating in the rise of self-reported stress levels and depressive symptoms [10]. What’s interesting to take into account is how many countries were implementing swift lockdown measures to mitigate the spread of COVID-19 [14]. In contrast, the United States of America (USA) opted for individual states to choose their lockdown policies. This move led to variation across the country. Mask policies varied from required to optional, businesses remained open or closed for different durations, and policymakers touted different overall messages. This made it difficult for individuals to determine what sources were correct and what information applied to the specific lockdown restriction they were in depending on their state. Timing of when a state enacted a mask mandate had a significant association with the spread of COVID-19 [14]. Recent research has suggested that state-level policy differences may be associated with different outcomes concerning the COVID-19 pandemic [37].

This disorganization from the federal government of the United States for individual states to handle their own lockdown issues was also prominent when it came to its vaccine roll-outs. When the Food and Drug Administration began vaccine rollout in December of 2020, there was a vacuum of leadership from the federal government in administrating vaccines to the population [15]. The previous administration provided little to no government insight on how states should run their vaccine roll-out, leaving most states to handle vaccine roll-out at their own discretion [15]. Moreover, vaccination sites were limited on how many shots they were able to administer, but there was a surplus of vaccines distributed to several states. The miscommunications between the federal government and the states caused many of these vaccines to not be given out to these important sites for the public [16]. The uneven vaccine roll-out to the public, spread of misinformation from the state and federal governments, as well as vaccine hesitancy has resulted in difficulty for the public to become vaccinated [17].

In the United States, prevalence of symptoms of anxiety disorder rose from the second quarter of 2019 (8.1%) to 25.5% in June 2020 [3]. Depressive symptoms also saw a reported increase from 6.5% (second quarter) to 24.3% (June 2020) [3]. We subsequently use the anxiety and depression as indicators of the mental health of the general population. Social isolation also affected the lifestyle and behavior of the public from sleep disorders, unhealthy eating habits, and restriction of in-person social activity, possibly leading to a further increase in mental health problems [7]. Social isolation runs the potential of being a risk factor for other problems such as dementia, premature death, and physiological distress [12]. The United States and many other countries at this stage of the pandemic experienced similar trends in their population from health care providers to the general public reporting elevated mental health issues likely stemming from the pandemic and lockdown restrictions [13]. Additionally, uneven vaccine roll-outs, COVID-19 incidence, and deaths throughout the timeline of COVID-19 in the United States have impacted the mental health of the public. State policies, geographic location, and political affiliation may be among some of the potential factors that can be used as mental health indicators.

At the same time, surveys have been issued across the population to ascertain the severity of the mental health problems of the population. This research aims to look at the presented data from COVID-19 Trends and Impact Surveys to assess the possible relationship between COVID-19 incidence and death and two social variables that were commonly impacted across the United States via lockdown policies with mental health indicators in each state in two time periods. Due to a change in the formatting of several questions, we had to split the analysis to accurately assess the survey results. The first time period ranges from September 9^*th*^, 2020 to March 2^*nd*^, 2021 and the second time period ranges from March 2^*nd*^, 2021 to January 10^*th*^, 2022. In this study, we use the aforementioned survey data collected in each state to assess the impact of both the spread of COVID-19 and the impact of reduced social contact as a result of the pandemic via variables related to social isolation on the mental health of the general population.

In Section 2, we provide further detail on the survey data analyzed and describe the formulation of models used for analysis. In Section 3, the coefficients of the fitted models and their impulse response functions are assessed. In the discussion section (Section 4), we present some implications of our results and frame them in the context of existing literature as well as presenting some limitations and strengths of the current study. Finally, in Section 5, we summarize our results and provide insight into future directions for this research.

## 2 Materials and Methods

### 2.1 Data

The data used in this study was collected and aggregated by the Delphi Research Group at Carnegie Mellon University in partnership with Facebook [18]. These surveys are issued by the Delphi Research Group at Carnegie Mellon which collects survey results about people’s responses to the current situation of the pandemic. These responses are distributed through the collaboration of Facebook and selected at random. The surveys themselves cover questions such as a person’s demographics, their mental health during the pandemic, how COVID-19 affected them, vaccine roll-out, and other important questions. These surveys allow investigators to compare responses across different regions of the United States and make informed public health decisions [18]. We used results from the COVID-19 Trends and Impact survey as well as case and death data provided by John Hopkins University, both of which can be accessed using the covidcast R package created by the Delphi Research Group. The survey asked respondents several questions regarding various topics, but we focus on the results of questions surrounding mental health and social distancing and travel for analysis. The survey results are weighted to be representative of the population of the United States and are reported as an estimated percentage.

As stated above, we perform our analysis on two subsets of the survey results due to the format of questions being changed on March 2^*nd*^ of 2021. Our first analysis covers the period of September 8^*th*^, 2020 to March 2^*nd*^, 2021 and the second analysis covers the period of March 2^*nd*^, 2021 to January 10^*th*^ 2022. As a measure of anxiety in the general population of each state, we used the ‘Estimated percentage of respondents who reported feeling nervous, anxious, or on edge for most or all of the past 5 days’. As a measure of depression, we used the ‘Estimated percentage of respondents who reported feeling depressed for most or all of the past 5 days’. We used two indicators as a proxy for social isolation. The first indicator was the ‘Estimated percentage of respondents who spent time with someone who isn’t currently staying with you in the past 24 hours’. The second indicator was the ‘Estimated percentage of respondents who worked or went to school outside their home in the past 24 hours’. This means that if an increase is observed in one of these variables, that is considered a decrease in the prevalence of social isolation in the general population. This also means that one might expect that these variables commonly have an inverse relationship with anxiety/depression (i.e. an increase in the percentage of respondents who worked or went to school should be associated with a decrease in anxiety/depression under normal circumstances).

In the context of the pandemic, there is a greater potential for the opposite effect to occur since individuals may be more worried about becoming sick with the more interactions they have. We used two measures of COVID-19 severity, the first being the number of new confirmed COVID-19 cases per 100,000 population per day and the second being the number of new confirmed deaths due to COVID-19 per 100,000 population per day. Due to the reporting schedule skipping weekends, we used the 7 day moving averages of these indicators to smooth the observed signals. As previously mentioned, the nature of questions changed slightly in the second period, for indicators used to measure anxiety and depression, the questions shifted from asking about the past 5 days to the past 7 days. For indicators used as a proxy for social isolation, the questions changed to ‘Estimated percentage of respondents who spent time *indoors* with someone who isn’t currently staying with you in the past 24 hours’ and ‘Estimated percentage of individuals who worked or went to school *indoors* and outside their home in the past 24 hours’. COVID-19 related indicators were unchanged in both time periods. The implications that these changes have will be discussed in Section 4. Models fit in the second time period also included the ‘Estimated percentage of respondents who have already received a vaccine for COVID-19’, and the results for that indicator are discussed in the Supplementary Figures. Next, we describe the methods used to analyze the survey data.

### 2.2 Model Formulation

The time series data described is non-stationary and co-integrated and thus was analyzed using Vector Error Correction Models (VECMs). To verify that our data were non-stationary and co-integrated, we ran an autocorrelation function and Johansen’s test for each state and found each time series to be non-stationary and co-integrated up to at least order 1 in every state. The results of each of these are provided in the results folder of the GitHub link (https://github.com/alxjfulk/social isolation and COVID19). VECMs are modified Vector Auto-Regressive Models (VARs) that are able to account for possible long-run relationships that arise in non-stationary, co-integrated time series data [19]. VARs have long been used to analyze economic data in an attempt to gain insight into what factors contribute to a particular economic variable [20] and we believe that the COVID-19 pandemic presents a unique opportunity to leverage those methods to gain insight into which factors (social isolation or COVID-19) seem to have more of an effect on the mental health of the general population of the United States. In order to make the results as interpretable as possible, we used the same general formula for each state with two lags of each variable included in each equation. From the urca package in R [21], a general VAR model is given as:

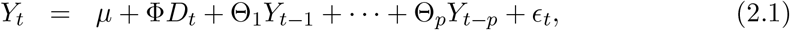

for *t* = 1, …, *T*. Where *μ* is a constant, *D*_*t*_ is the trend matrix, Θ_*i*_ is the coefficient matrix, *ϵ*_*t*_ is the error vector, and *Y*_*t−i*_ is the vector of variables. Then, our VECM is specified as:

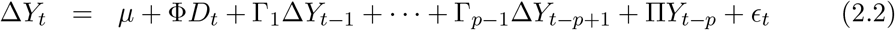

with:

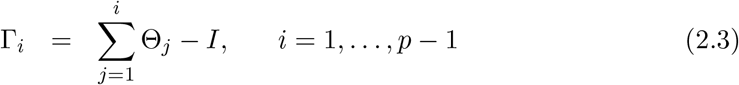

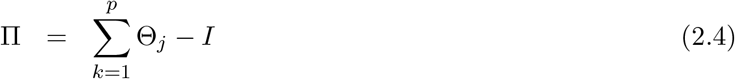

Where the Γ_*i*_ matrices contain cumulative long-run impacts of variables. Notably, since point estimates of each variable can be biased as mentioned in the limitations section of the survey data, we evaluate the sign (positive, negative, or not significant) of each variable in the equations for anxiety and depression in each state as the surveys are likely to effectively capture when a variable increases or decreases. This allows us to make inferences on the effect of each variable on anxiety and depression without actually having to evaluate the estimates themselves, thus limiting the effect of that bias. In addition, we converted each VECM into a VAR using the vec2var function in the vars package for the purpose of estimating impulse response functions (IRFs) for each variable of interest [22]. Our VECM equations are given in Appendix A.

## 3 Results

### 3.1 Vector Error Correction Model Results

For the sake of brevity, we only present the results of the VECMs fitted to state data in the time period from March 2^*nd*^, 2021 to January 10^*th*^, 2022. The results of the VECMs fitted using data from the time period of September 8^*th*^, 2020 to March 2^*nd*^, 2021 are provided in the Appendix B. Furthermore, the results of the VECMs fitted using data on the percentage of individuals vaccinated in the second time period are given in Appendix C. Note that we abbreviate the variables related to social isolation (i.e. ‘estimated percentage of respondents who spent time indoors with someone who isn’t currently staying with you in the past 24 hours’ and ‘estimated percentage of individuals who worked or went to school indoors and outside their home in the past 24 hours’) as time spent w/ others and work outside home, respectively.

#### 3.1.1 Effects of COVID-19 in the Second Time Period

Figure 1 shows which states had positive, negative, or non-significant VECM coefficients for COVID-19 incidence and death in the second time period. A total of 30 states returned significant positive coefficients and one state returned significant negative coefficients for COVID-19 incidence, indicating that as incidence increases, so does anxiety in the general population. This does not mean that COVID-19 is directly causing this increase, though it may be to some degree based on previous studies [10]. Rather, the many factors surrounding an increase in COVID-19 incidence, such as possible school and work closing, increased limits on gatherings, and media coverage of the topic, are likely contributing to this increase in anxiety. We see much less of an impact of COVID-19 related death on anxiety, however, there are still several significant coefficients. We see mostly positive results with 11 states returning positive coefficients and 3 states returning significant negative coefficients. There are many more positive results in the former case when comparing the impact of COVID-19 incidence versus death. This may mean that people are overall more concerned with becoming infected with the virus than dying from it.

**Figure 1:**
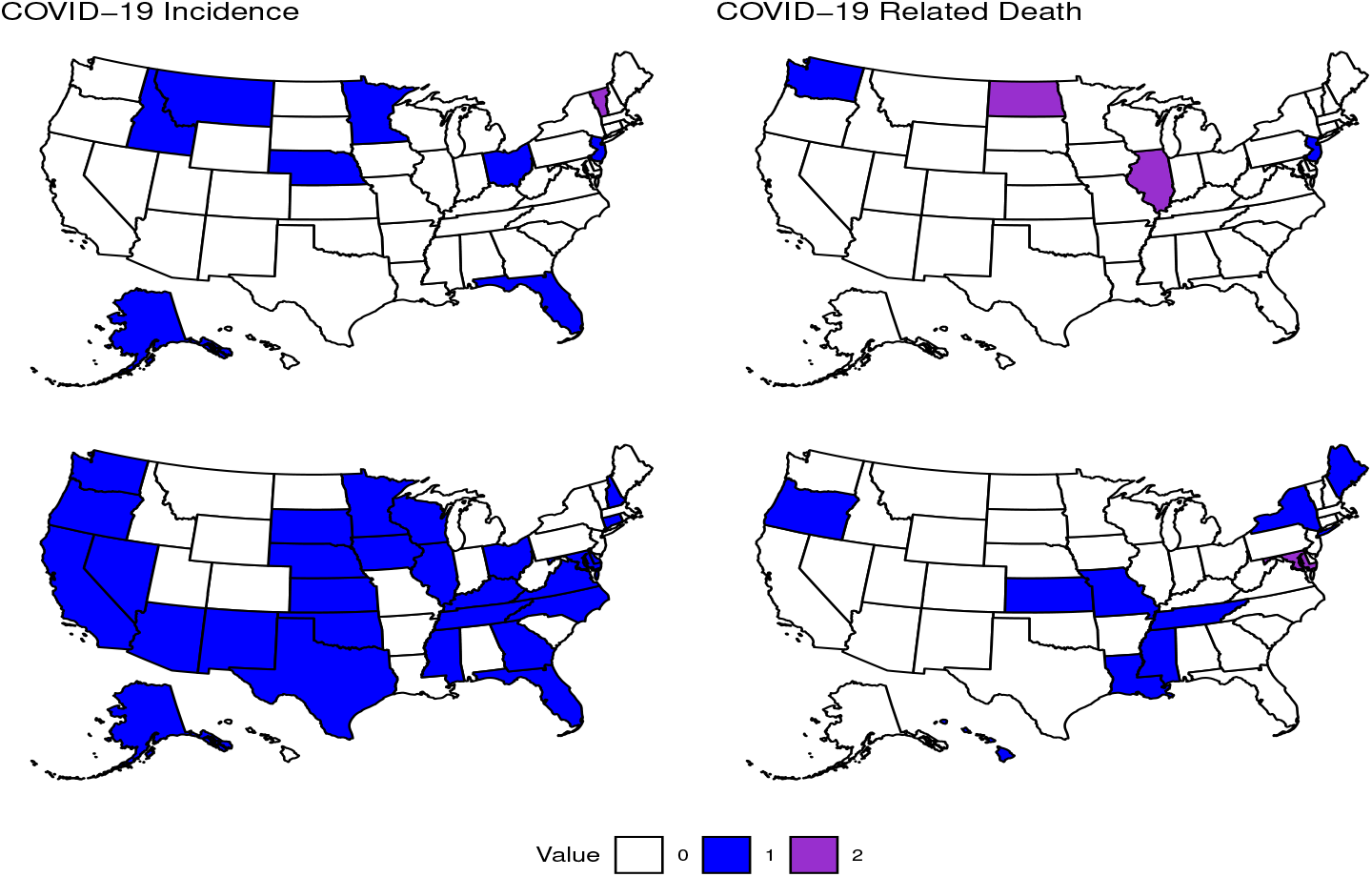
Effect of COVID-19 on Anxiety. States that had significantly positive or negative coefficients in their equation for anxiety for the first or second lag of variables related to COVID-19 in the second time period. The color depicts whether a state had a positive (blue), negative (purple), or no significant coefficient for the corresponding variable. COVID-19 Incidence lag 1 (top left): (positive) Alaska, Florida, Idaho, Minnesota, Montana, Nebraska, New Jersey, (negative) Vermont. Lag 2 (bottom left): (positive) Alaska, Arizona, California, Connecticut, Delaware, Florida, Georgia, Iowa, Illinois, Kansas, Kentucky, Maryland, Minnesota, Mississippi, Nebraska, Nevada, New Hampshire, New Jersey, North Carolina, Ohio, Oklahoma, Oregon, South Dakota, Tennessee, Texas, Virginia, Washington, Wisconsin. COVID-19 Related Death lag 1 (top right): (positive) New Jersey, Washington, (negative) Illinois, North Dakota. Lag 2 (bottom right): (positive) Hawaii, Kansas, Louisiana, Maine, Mississippi, Missouri, New York, Oregon, Tennessee, (negative) Maryland.

Thirteen states had significant positive coefficients for COVID-19 incidence and one state returned a negative coefficient in their equations for depression as shown in Figure 2. COVID-19 related death had 12 states with positive coefficients and two states with negative coefficients. Here we also see mostly positive results, however there seems to be less of an impact of these variables on depression compared to anxiety, though it is possible that we were not able to account for enough lags to see the longer-term effects of these variables. Notice that, while rare and only for the first lag of COVID-19 related death, there are some states that had significant coefficients for depression, but not anxiety (Iowa, Nevada, New Jersey, and West Virginia).

**Figure 2:**
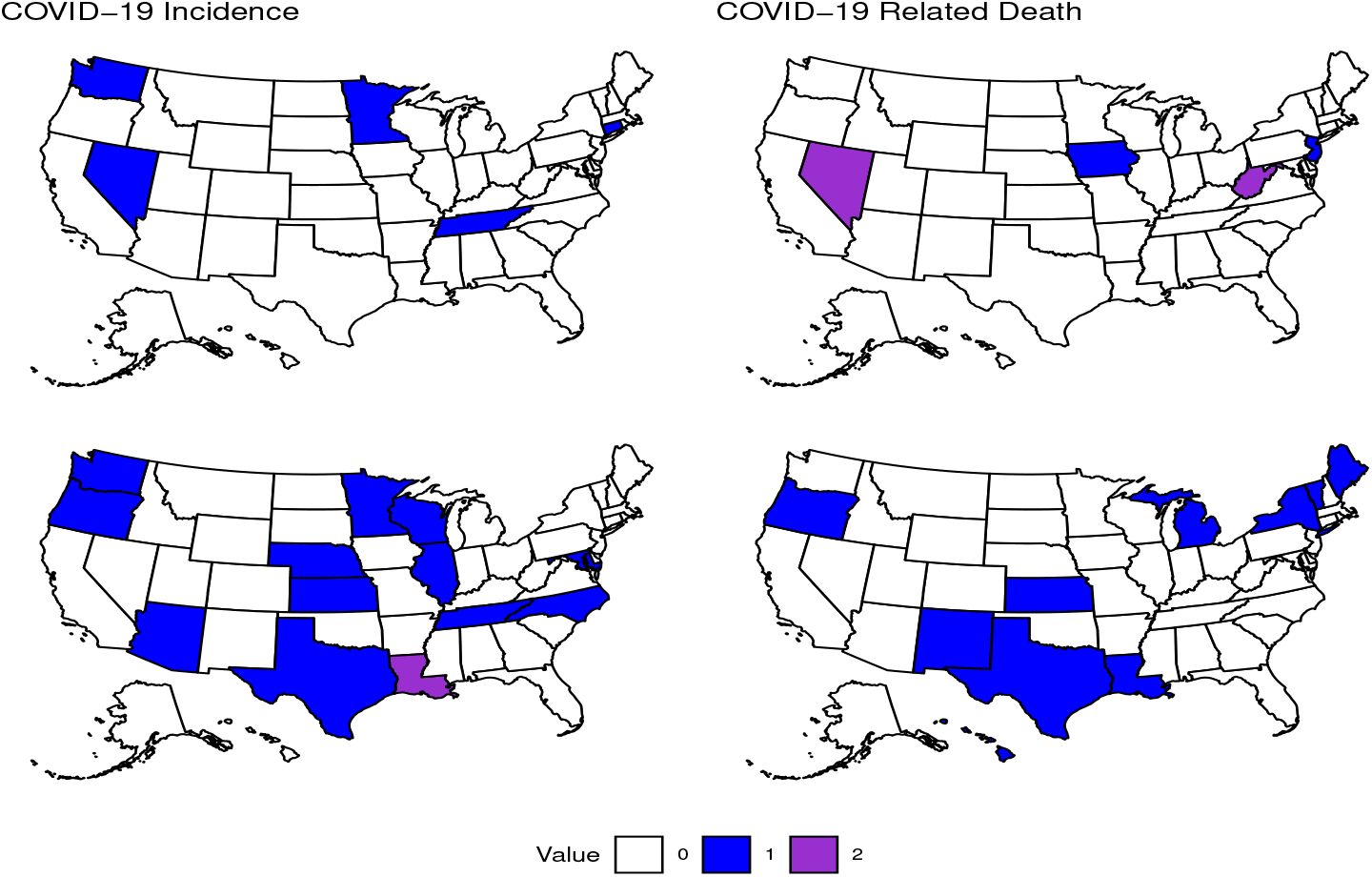
Effect of COVID-19 on Depression. States that had significantly positive or negative coefficients in their equation for depression for the first or second lag of variables related to COVID-19 in the second time period. The color depicts whether a state had a positive (blue), negative (purple), or no significant coefficient for the corresponding variable. COVID-19 Incidence lag 1: (positive) Connecticut, Minnesota, Nevada, Tennessee, Washington. Lag 2: (positive) Arizona, Illinois, Kansas, Maryland, Minnesota, Nebraska, North Carolina, Oregon, Washington, Wisconsin, (negative) Louisiana. COVID-19 Related Death lag 1: (positive) Iowa, New Jersey, (negative) Nevada, West Virginia. Lag 2: (positive) Hawaii, Kansas, Louisiana, Maine, Michigan, New Mexico, New York, Oregon, Texas, Vermont.

#### 3.1.2 Effects of Social Isolation in the Second Time Period

Figure 3 shows that time spent with others indoors had a total of five states with significant positive coefficients for this variable and five states with significant negative coefficients. No states in the western United States returned significant results for this variable. There is also a break in the pattern of more significant results in the second lag with time spent with others indoors and there is no clear indication as to whether this variable is associate more with an increase or decrease in anxiety. Working outside of the home indoors gave significant positive coefficients for nine states and negative coefficients for five states. The results were more varied for this variable in terms of geographic distribution compared to time spent with others indoors. It does tend to happen that an increase in people working outside their homes indoors is associate with an increase in anxiety, but it is not a strict rule as more than 35% of the total number of significant states for this variable had negative coefficients.

**Figure 3:**
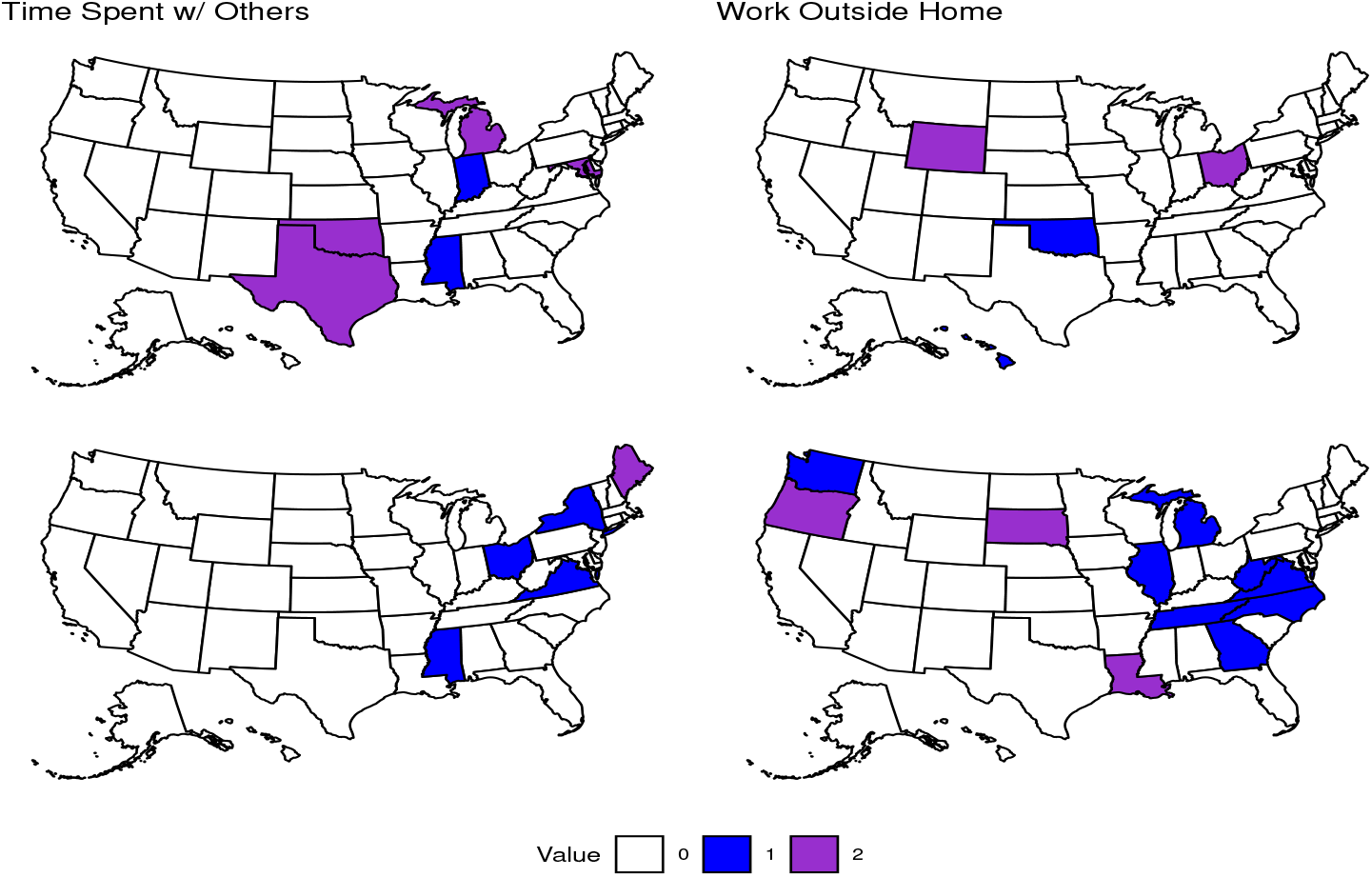
Effect of Social Isolation on Anxiety. States that had signifcantly positive or negative coefficients in their equation for anxiety for the first or second lag of variables related to Social Isolation in the second time period. The color depicts whether a state had a positive (blue), negative (purple), or no significant coefficient for the corresponding variable. Time Spent w/ Others lag 1: (positive) Indiana, Mississippi, (negative) Maryland, Michigan, Oklahoma, Texas. Lag 2: (positive) Mississippi, New York, Ohio, Virginia, (negative) Maine. Work Outside Home lag 1: (positive) Oklahoma, (negative) Ohio, Wyoming. Lag 2: (positive) Georgia, Illinois, Michigan, North Carolina, Tennessee, Virginia, Washington, West Virginia, (negative) Louisiana, Oregon, South Dakota.

Finally, we describe the number of states with significant coefficients for social variables in the second time period in equations for depression (see Figure 4). Nineteen states returned positive results and four states returned negative results for the variable time spent with others indoors. Note that one state (Michigan) had a negative coefficient for the first lag and a positive coefficient for the second lag of this variable, making it difficult to determine the actual effect it had on the population in that state. The variable estimating the percentage of individuals working outside their home indoors had nine states with positive results and nine states with negative coefficients. In this case, two states went from negative in the first lag to positive in the second lag. Here, we see a slight preference for more (positive) results in the second lag of the first variable and a slight preference for negative results in the first lag and positive results in the second lag of the second variable. Unlike what was observed in the variables related to COVID-19, there is much less of an association between which states were significant for anxiety versus depression. Overall, this can be interpreted as variables related to COVID-19 having a much more predictable and mostly positive effect on the prevalence of mental health issues whereas since the interactions caused by an increase in social variables can be good or bad, these variables were less significant and less reliable in terms of their affect the prevalence of anxiety and depression. For both anxiety and depression, we have either more or an equal number of states with significant coefficients in the second lag of variables compared to the first. This preference for later lags is also evident in the results for the first time period as well and it may indicate that it is important to evaluate more lags if possible when evaluating the effects of certain variables on anxiety and depression.

**Figure 4:**
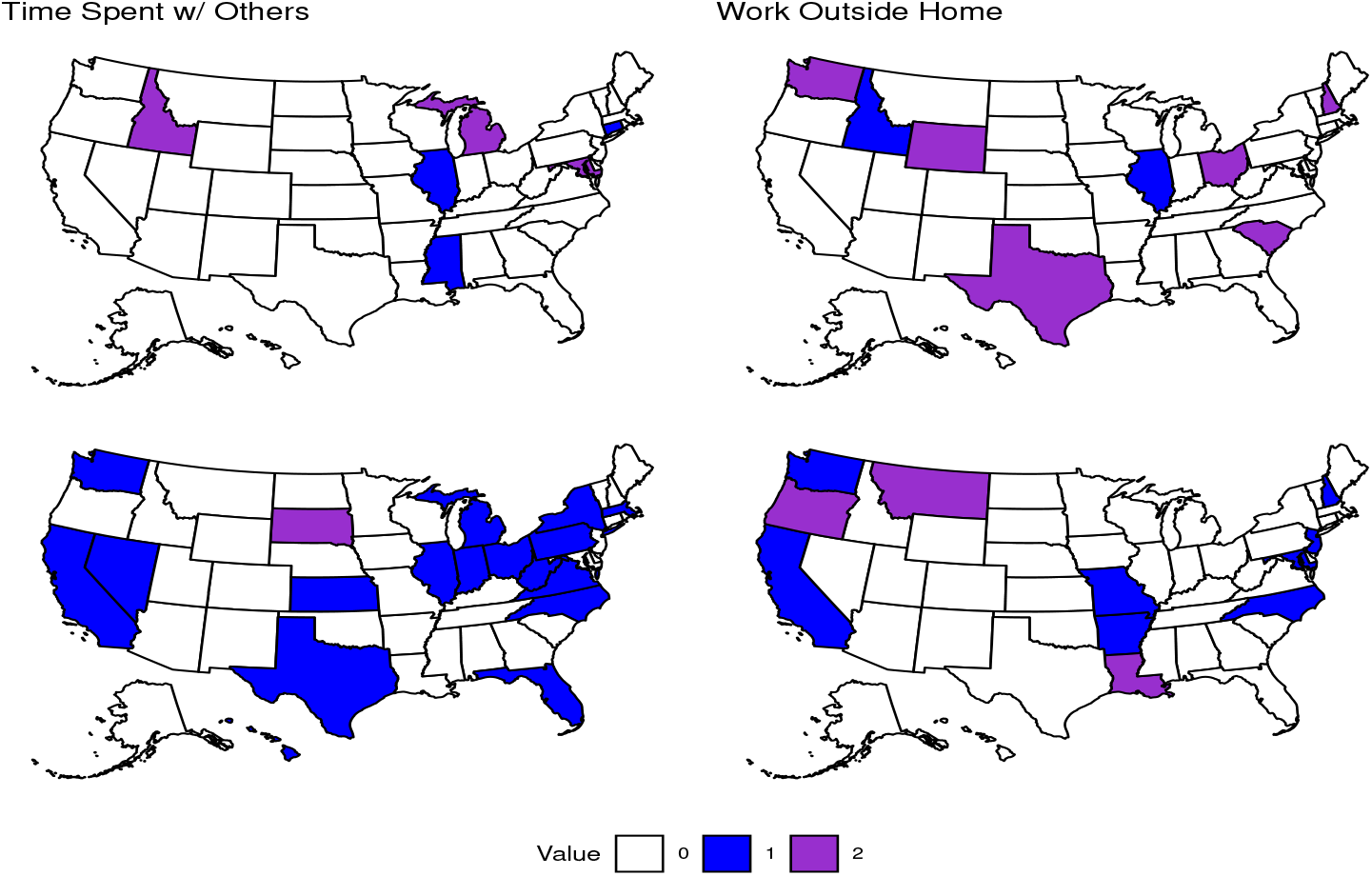
Effect of Social Isolation on Depression. States that had significantly positive or negative coefficients in their equation for depression for the first or second lag of variables related to Social Isolation in the second time period. The color depicts whether a state had a positive (blue), negative (purple), or no significant coefficient for the corresponding variable. Time Spent w/ Others lag 1: (positive) Connecticut, Illinois, Mississippi, (negative) Idaho, Maryland, Michigan. Lag 2: (positive) California, Florida, Hawaii, Illinois, Indiana, Kansas, Massachusetts, Michigan, Nevada, New York, North Carolina, Ohio, Pennsylvania, Texas, Virginia, Washington, West Virginia, (negative) South Dakota. Work Outside Home lag 1: (positive) Idaho, Illinois, (negative) New Hampshire, Ohio, South Carolina, Texas, Washington, Wyoming. Lag 2: (positive) Arkansas, California, Maryland, Missouri, New Jersey, North Carolina, Washington, (negative) Louisiana, Montana, Oregon.

### 3.2 Impulse Response Function Results

Due to the nature of impulse response functions (IRFs), we are able to more easily interpret the effect that an increase in each variable has on anxiety and depression in the general population. Again, seeing a significant result for one variable after increasing another does not imply that the latter variable is causing the increase in the former. Likely due to the fact that we have fewer data points in the first time period, there are more differences between significant results in the first time period’s IRFs compared to the coefficient results discussed previously. Due to that and for the sake of brevity, we present only the results for the second time period and provide the results of the first time period as well as the IRF results from the percentage of individuals vaccinated in the second time period in the Appendices B.3 and Appendix C.

#### Effects of COVID-19 in the Second Time Period

For the effect of an impulse of COVID-19 incidence on the prevalence of anxiety, we have a similar behavior as when we were looking at coefficients as there are 13 states that increased in anxiety and 3 states that decreased following an increase in incidence as shown in Figure 5. An important thing to notice here is that there are fewer states with significant results compared to the coefficients of the first and second lag of this variable. This may indicate that one should consider more aspects than just the IRF when analyzing the effects of different variables in a VAR or VECM. Moving on to COVID-19 related death, there were 4 states that returned significant positive responses and 6 states that returned significant negative responses. In this case, there is a clear difference in the geographic distribution of significant impulses compared to the coefficient results, further highlighting the need for in-depth analysis of each aspect of our models. Since there are 5 states that have significant impulses that did not have significant coefficients for either the first or second lag of COVID-19 related death, it may be that there are more intricate relationships going on between variables than simply that an increase in one causes an increase in another. More likely is that an increase in COVID-19 related death leads to or coincides with changes in other variables that then have a more significant impact on anxiety in a particular state.

**Figure 5:**
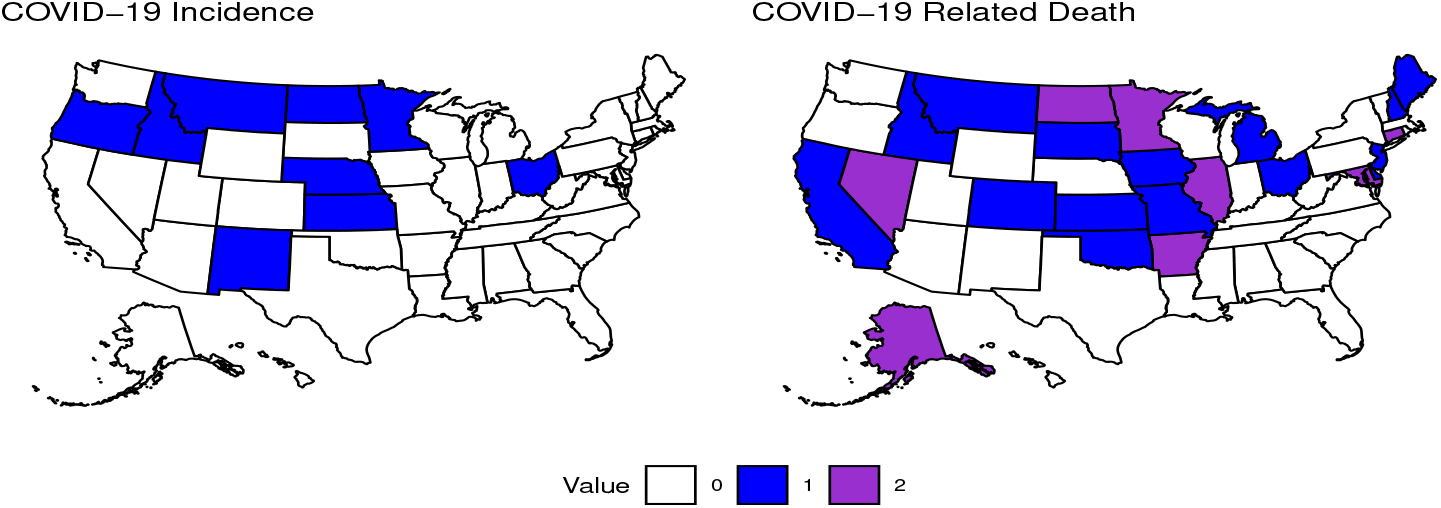
Effect of COVID-19 on Anxiety. States that had significantly positive or negative impulses for anxiety for variables related to COVID-19 in the second time period. The color depicts whether a state had a positive (blue), negative (purple), or no significant coefficient for the corresponding variable. COVID-19 Incidence (left): (positive) Alaska, Arizona, Arkansas, Florida, Hawaii, Idaho, Kansas, Minnesota, Montana, Nebraska, Nevada, Ohio, Oregon, (negative) North Dakota, Vermont, Wisconsin. COVID-19 Related Death (right): (positive) Idaho, Maine, Missouri, Ohio, (negative) Arizona, Arkansas, Illinois, Nevada, North Dakota, Oregon.

The results of the IRFs of COVID-19 related variables on depression are presented in Figure 6 and they paint a similar picture to the coefficient results, though there are some exceptions as was the case with the IRFs for anxiety. An increase in COVID-19 incidence indicated a positive impulse in depression for 8 states and a negative impulse for 2 states. So, we again see that an increase in COVID-19 incidence is associated with an increase in anxiety. An impulse in COVID-19 related death led to a positive impulse in depression for 8 states and a negative impulse for 3 states. There are also several states that have significant coefficients for COVID-19 incidence or COVID-19 related death that did not have significant impulses and vice versa.

**Figure 6:**
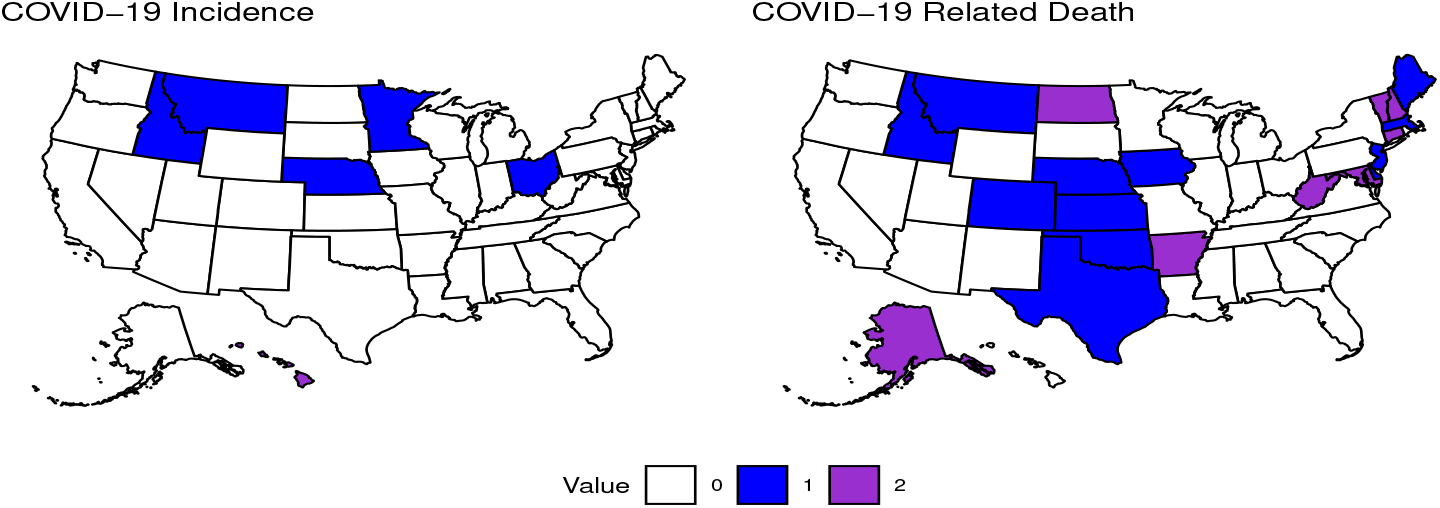
Effect of COVID-19 on Depression. States that had significantly positive or negative impulses for depression for variables related to COVID-19 in the second time period. The color depicts whether a state had a positive (blue), negative (purple), or no significant coefficient for the corresponding variable. COVID-19 Incidence (left): (positive) Arizona, Kansas, North Dakota, Ohio, Oregon, (negative) Hawaii, New Hampshire. COVID-19 Related Death (right): (positive) Delaware, Idaho, Maine, Minnesota, Ne-braska, Nevada, New Hampshire, New Jersey, Oklahoma, Tennessee, Texas, (negative) Arizona, Oregon, West Virginia.

While the results of the IRFs for the effect of social variables on anxiety show a similar number of significant states compared to the coefficient results (11 vs 10), there is a notable difference in which states are significant in each case (see Figures 3 and 7). There are 6 states that had a positive impulse following an impulse in time spent with others indoors and 5 states with a negative impulse. The effect of working outside the home indoors had a clearer split in that only 3 states had a positive impulse and 10 states had a negative impulse. The variable time spent with others indoors likely captures some interactions with individuals that the respondent isn’t familiar with which may explain why we commonly see positive impulses in anxiety following an impulse in this variable. Another possibility is that this variable does not provide any insight into whether the time spent with someone a respondent isn’t currently staying with is positive or negative, so there may be negative social interactions that are not directly linked to a respondent being worried about contracting COVID-19. On the other hand, there are many more states that had significant negative impulses for working outside the home indoors. While this variable also does not capture whether a respondent has a positive or negative work environment, the benefits of consistency and stability of in-person employment may outweigh the negative effects of a poor work environment.

**Figure 7:**
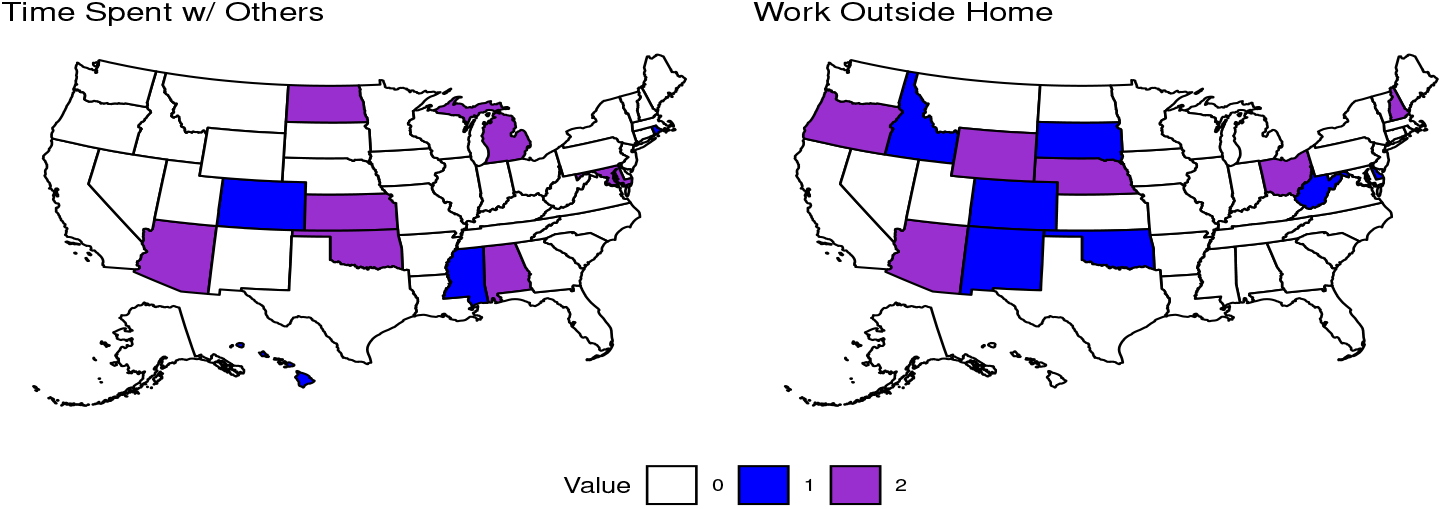
Effect of Social Variables on Anxiety. States that had significantly positive or negative impulses for anxiety for social variables in the second time period. The color depicts whether a state had a positive (blue), negative (purple), or no significant coefficient for the corresponding variable. Time spent with others indoors (left): (positive) Arkansas, Florida, Hawaii, Indiana, Mississippi, Nebraska, (negative) Arizona, Kansas, Michigan, North Carolina, Oklahoma.Work outside home indoors (right): (positive) New Mexico, North Dakota, Oklahoma, (negative) Arizona, Florida, Hawaii, Louisiana, Nebraska, Nevada, New Hampshire, Ohio, Oregon, Wyoming.

The IRF results for the variables related to social isolation are given in Figure 8. Similar to the effects that social variables had on anxiety, we see a roughly even split in terms of significant impulses for time spent with others indoors with 5 states have positive impulses and 4 states having negative impulses. For working outside the home indoors, there were 4 states with positive impulses and 8 states with negative impulses.

**Figure 8:**
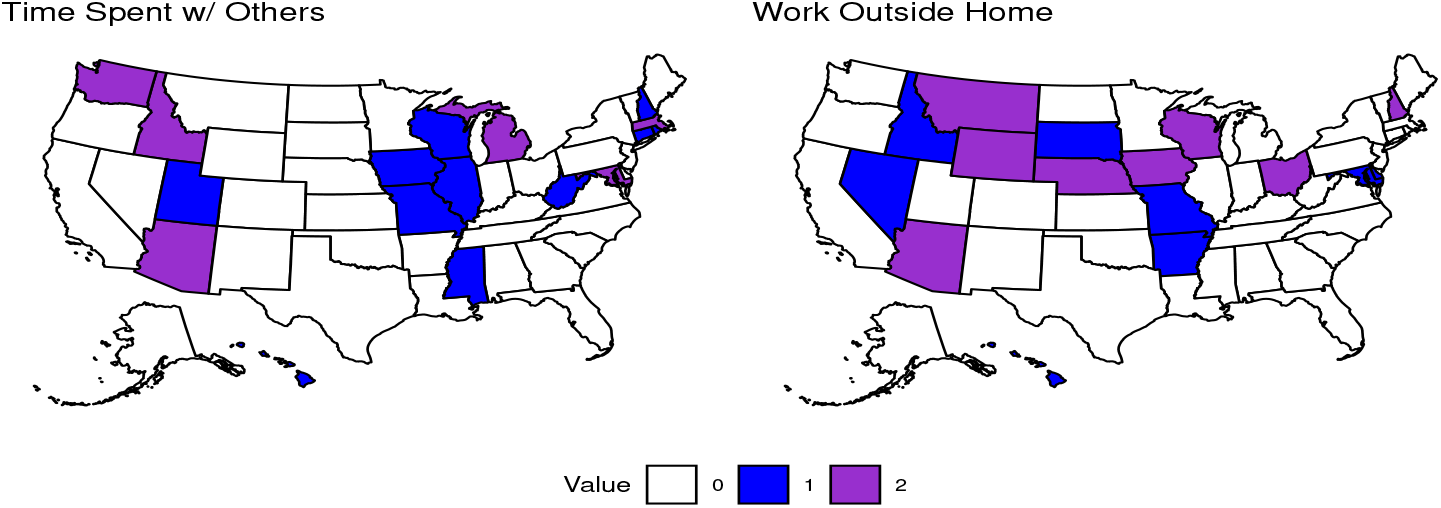
Effect of Social Variables on Depression. States that had significantly positive or negative impulses for anxiety for social variables in the second time period. The color depicts whether a state had a positive (blue), negative (purple), or no significant coefficient for the corresponding variable. Time spent with others indoors (left): (positive) Arkansas, Illinois, Mississippi, Ohio, Wisconsin, (negative) Arizona, Idaho, Maryland, Michigan. Work outside home indoors (right): (positive) Delaware, Maryland, Nevada, South Dakota, Wisconsin, (negative) Arizona, Iowa, Nebraska, New Hampshire, Ohio, Oregon, Wisconsin, Wyoming.

## 4 Discussion

The results of the impacts of COVID-19 on mental health in the general population have two major highlights. First, we see that COVID-19 incidence and death played significant roles in determining the prevalence of anxiety and depression in both time periods (see Figures 1, 2, 5, 6 and Appendix B for results in the first time period). When not double counting states that had the same sign of significant results in each lag, we had a total of 74 significant positive coefficients and 3 significant negative coefficients for the COVID-19 incidence variable across both time periods. COVID-19 related death had 25 significant positive coefficients returned and 13 significant negative coefficients across both time periods. Thus, for a majority of states, when we see a rise in COVID-19 incidence or death, we also tended to see an increase in the prevalence of anxiety and depression in the general population. This trend held for each variable in each time period except for COVID-19 related death in the first time period. It is possible that respondents were more concerned with being infected with the virus than actually dying of COVID-19 during this time period. These results are in line with previous studies that have provided some insight into possible links between COVID-19 infection and death and development of depressive symptoms and they are important to consider in the context of future pandemics. The observed association between these variables may have been enhanced by external factors like the influence of media [34, 36, 28].

Several past studies have unearthed possible links between COVID-19 infection and development of anxious or depressive symptoms [24, 25, 26, 27, 29, 30, 31, 32, 28, 23] and while our study does not address that specific topic, it does imply that an increase in COVID-19 incidence and death may lead to increased anxiety and depression and this can be exacerbated by factors that coincide with increased prevalence of disease such as continuous media coverage and the possibility of increased medical bills and loss of wages due to long-term issues that can arise with COVID-19 infection [33, 34]. Second, variables related to social isolation did have a significant impact on anxiety and depression in the general population (see Figures 3, 4, 7, and 8), but perhaps less so than the impact that variables related to COVID-19 had. We had a total of 41 significant positive coefficients and 16 significant negative coefficients for individuals who spent time indoors with others they aren’t currently staying with. For individuals working outside their home, we calculated 27 significant positive coefficients and 22 significant negative coefficients. In the case of our variables related to social isolation, we see significant variation across both time periods and although there are fewer significant coefficients for these variables, the negative impacts of social isolation are clear [34, 23] and some effort should be put forth to minimize their impacts.

There are some limitations and strengths of the study presented that should be noted. First, some limitations include that the questions used as proxies for social isolation (i.e. ‘estimated percentage of respondents who spent time indoors with someone who isn’t currently staying with you in the past 24 hours’ and ‘estimated percentage of individuals who worked or went to school indoors and outside their home in the past 24 hours’) likely do not accurately reflect the percentage of respondents who felt socially isolated. This is because of the fact that one can have social interaction and still feel socially isolated and also because individuals may be interacting online rather than in-person. Other limitations have to do with the survey data itself, namely the survey data used to fit models for each state was weighted representative of each state’s age and gender demographics, but states may differ in other demographics that are not accounted for. Additionally, there is limited data for some regions in states as regions that had less than 100 responses for a particular question were removed from the overall results of a state. Finally, the change in the wording of the questions from the first time period to the second likely changed how respondents interpreted the questions and may have impacted the results. There are also some strengths of the current study that should be highlighted. First, we have used a robust analytical framework for the analysis of this survey data that allows insight into mechanisms that impacted the prevalence of anxiety and depression in the United States across the studied period. Second, our measures for COVID-19 incidence and death are standard and robust, leaving little room for error in interpretation of results regarding those indicators.

## 5 Conclusions

The effects of the COVID-19 pandemic are far reaching and not fully understood, especially in terms of the effects that it has had on the mental health of the general population. This study used an approach commonly employed in economics, namely multiple time-series analysis using vector error-correction models, in an attempt to gain insight into the most significant factors surrounding the pandemic during two time periods ranging from September 8^*th*^, 2020 to March 2^*nd*^, 2021 and March 2^*nd*^, 2021 to January 10^*th*^, 2022, respectively. We found that indicators related to COVID-19 incidence and death commonly led to an increase in anxiety and depression in the general population across both studied periods. Indicators related to social isolation did not have a clear interpretation as there was significant variation across time and space. It is our hope that this paper provides some insight and guidance into how each state and business should tailor policies to their respective populations to minimize the effects of the variables included in this study on the mental health of those populations. In future research, we will use network analysis with these time series data to gain insights about how the states are clustered together with respect to a particular indicator as well as which policies had significant impacts on those indicators. We will then compare the results obtained with findings from this study.

## Data Availability

Data used are available at the link https://doi.org/10.1101/2021.07.24.21261076
All codes, data and results are available at the GitHub link: https://github.com/alxjfulk/social_isolation_and_COVID19

## Data Availability

Data used are available at the link https://doi.org/10.1073/pnas.2111452118

All code, data, and results are available at the GitHub link: https://github.com/alxjfulk/social_isolation_and_COVID19

## Author contributions

Alexander Fulk: Conceptualization, formal analysis, writing - original draft preparation, reviewing and editing; Raul Saenz-Escarcega: Conceptualization, writing - review, Hiroko Kobayahsi: Conceptualization, writing - review, and editing; Innocent Maposa: Formal analysis, writing - review, Folashade B. Agusto: Conceptualization, project administration, supervision, writing-reviewing and editing, funding acquisition.

## Acknowledgement

This research is supported by National Science Foundation under the grant number DMS 2028297.

## A VECM Equation Example

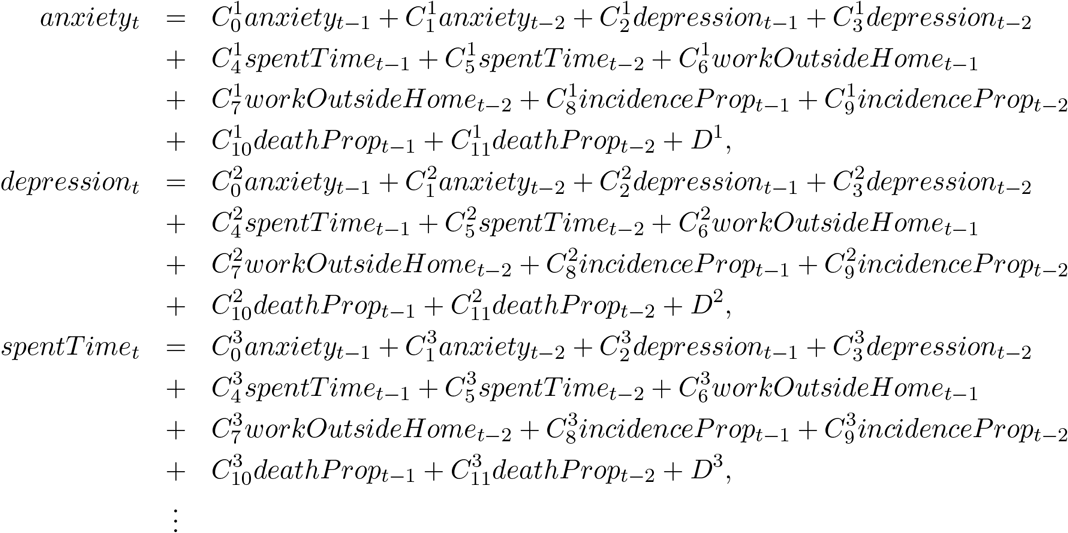

The equations for the other variables of each VECM follow a similar format. Table 1 provides a brief description of each variable listed in the equations above.

**Table 1:** Description of the variables for model described above.

| Variable | Meaning |
| --- | --- |
| *anxiety* | Estimated percentage of respondents who reported feeling nervous, anxious, or on edge for most or all of the past 5 days. |
| *depression* | Estimated percentage of respondents who reported feeling depressed for most or all of the past 5 days. |
| *spentTime* | Estimated percentage of respondents who spent time with someone who isn't currently staying with you in the past 24 hours. |
| *workOutsideHome* | Estimated percentage of respondents who worked or went to school outside their home in the past 24 hours. |
| *incidenceProp* | Number of new confirmed COVID-19 cases per 100,000 population per day. |
| *deathProp* | number of new confirmed COVID-19 deaths per 100,000 population per day. |
| $C_j^i$ | $j^{th}$ coefficient of the $i^{th}$ equation. |
| $D^i$ | constant associated with the $i^{th}$ equation. |

## B Results of the First Time Period

### B.1 Effects of COVID-19 in the First Time Period

**Figure 9:**
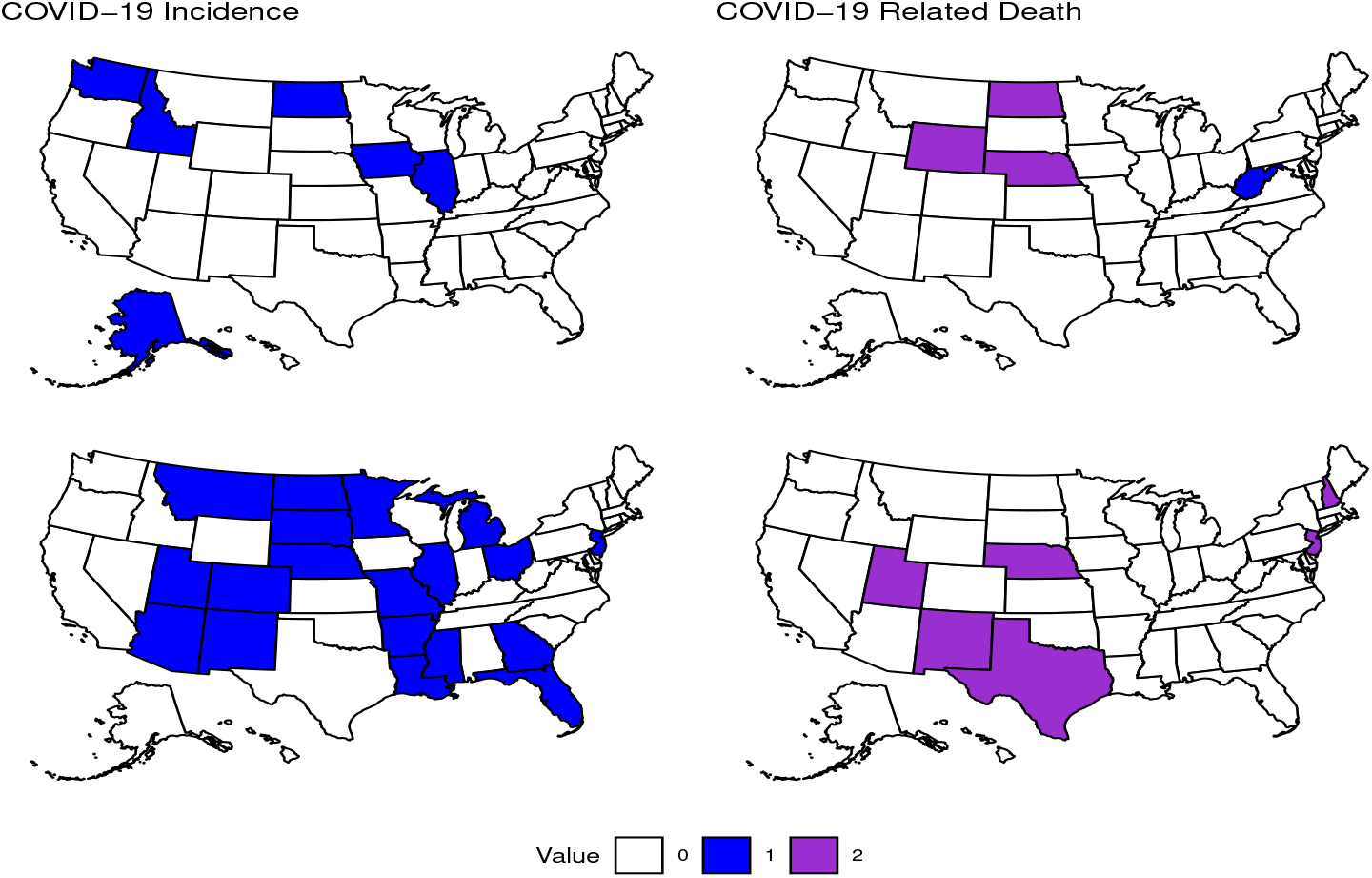
Effect of COVID-19 on Anxiety. States that had significantly positive or negative coefficients in their equation for anxiety for the first or second lag of variables related to COVID-19 in the first time period. The color depicts whether a state had a positive (blue), negative (purple), or no significant coefficient for the corresponding variable. COVID-19 Incidence lag 1: (positive) Alaska, Idaho, Illinios, Iowa, North Dakota, Washington, (negative) NA. Lag 2: (positive) Arizona, Arkansas, Colorado, Florida, Georgia, Illinois, Louisiana, Michigan, Minnesota, Mississippi, Missouri, Montana, Nebraska, New Jersey, New Mexico, North Dakota, Ohio, South Dakota, Utah, (negative) NA. COVID-19 Death lag 1: (positive) West Virginia, (negative) Nebraska, North Dakota, Wyoming. Lag 2: (positive) NA, (negative) Nebraska, New Jersey, New Mexico, Texas, Utah, Vermont

Twenty-three states had positive coefficients for COVID-19 incidence and all of these states returned positive values, indicating that an increase in incidence led to an increase in anxiety during the first time period. In contrast, almost all states that returned significant coefficients for COVID-19 related death had negative values and this variable had a much smaller impact overall; One state returned a positive coefficient and 8 states returned negative coefficients for this variable. Lastly, in both variables, we see more significant results in the second lag, which alludes to possible longer-term impacts of COVID-19 on anxiety and depression.

**Figure 10:**
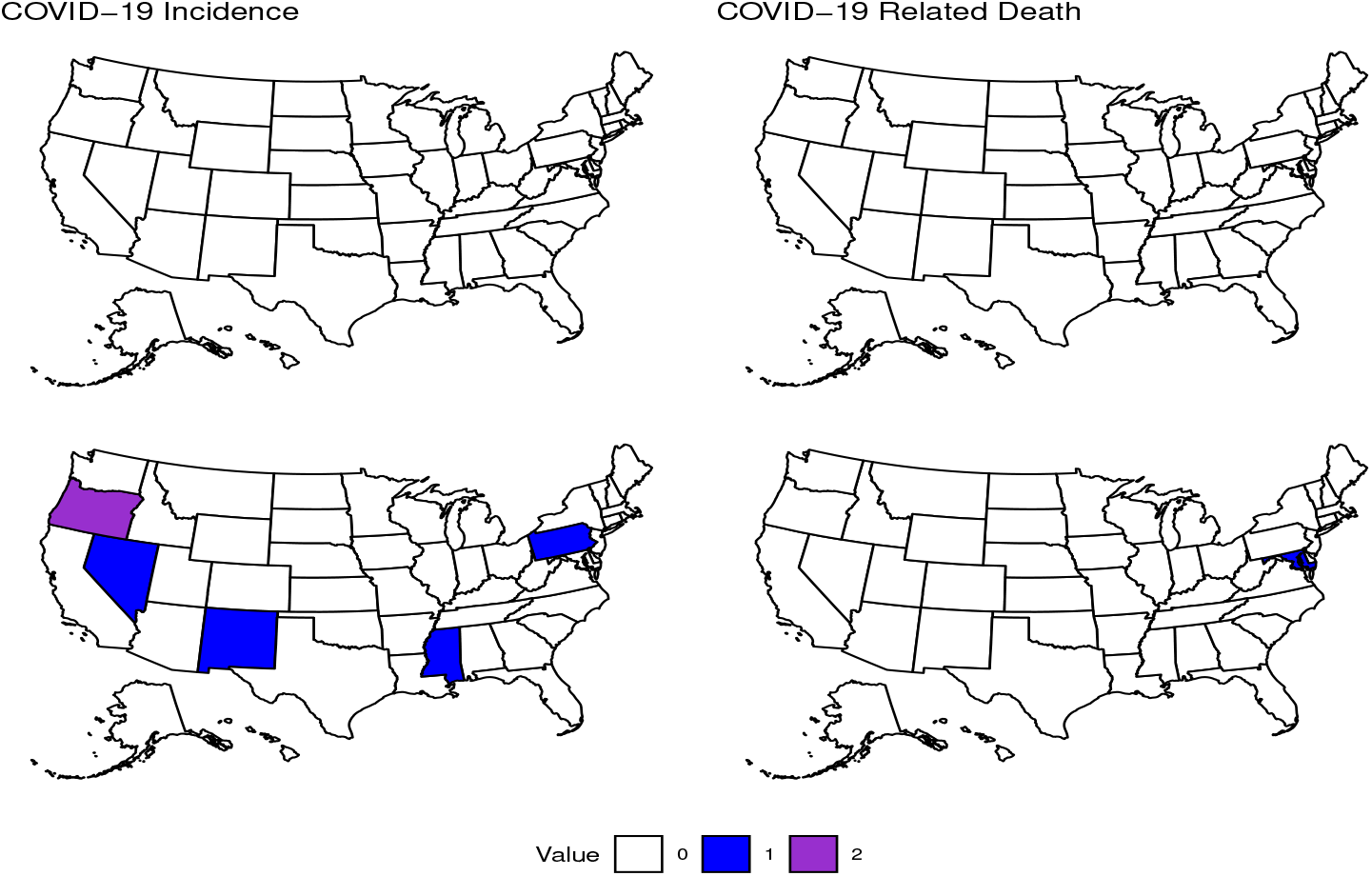
Effect of COVID-19 on Depression. States that had significantly positive or negative coefficients in their equation for depression for the first or second lag of variables related to COVID-19 in the first time period. The color depicts whether a state had a positive (blue), negative (purple), or no significant coefficient for the corresponding variable. COVID-19 Incidence lag 2: (positive) Pennsylvania, Mississippi, New Mexico, Nevada, (negative) Oregon. COVID-19 death lag 2: (positive) Maryland, (negative) NA.

Only a handful of returned significant coefficients when analyzing the impact that COVID-19 may have on depression in the first time period. When looking at COVID-19 incidence, we see that 4 states returned a positive coefficient for this variable and 1 state returned a negative coefficient for this variable. COVID-19 related death only had one state return a significant positive result. Due to the lack of significant coefficients for these variables, it is difficult to make inferences, but there are several reasons why this may have occurred. First, it may be that we do not have enough data points to capture possible relationships among depression and our COVID-19 related variables. Second, the structure of our model may be ill-suited to capturing possible relationships between these variables. We chose to only include two lags of each variables, but it may be that we need to include more lags to assess the impacts that occur over a longer period of time. This is somewhat supported in our results since the second lag of each variable usually contains more significant coefficients. Finally, it may be that there was little impact of COVID-19 on depression in this time period. With the one state that returned significant results, we again see that the second lags of each contain the most significant results, further supporting the idea that the impacts of these variables on anxiety and depression take time to reveal themselves at the level of the general population.

### B.2 Effects of Social Isolation in the First Time Period

**Figure 11:**
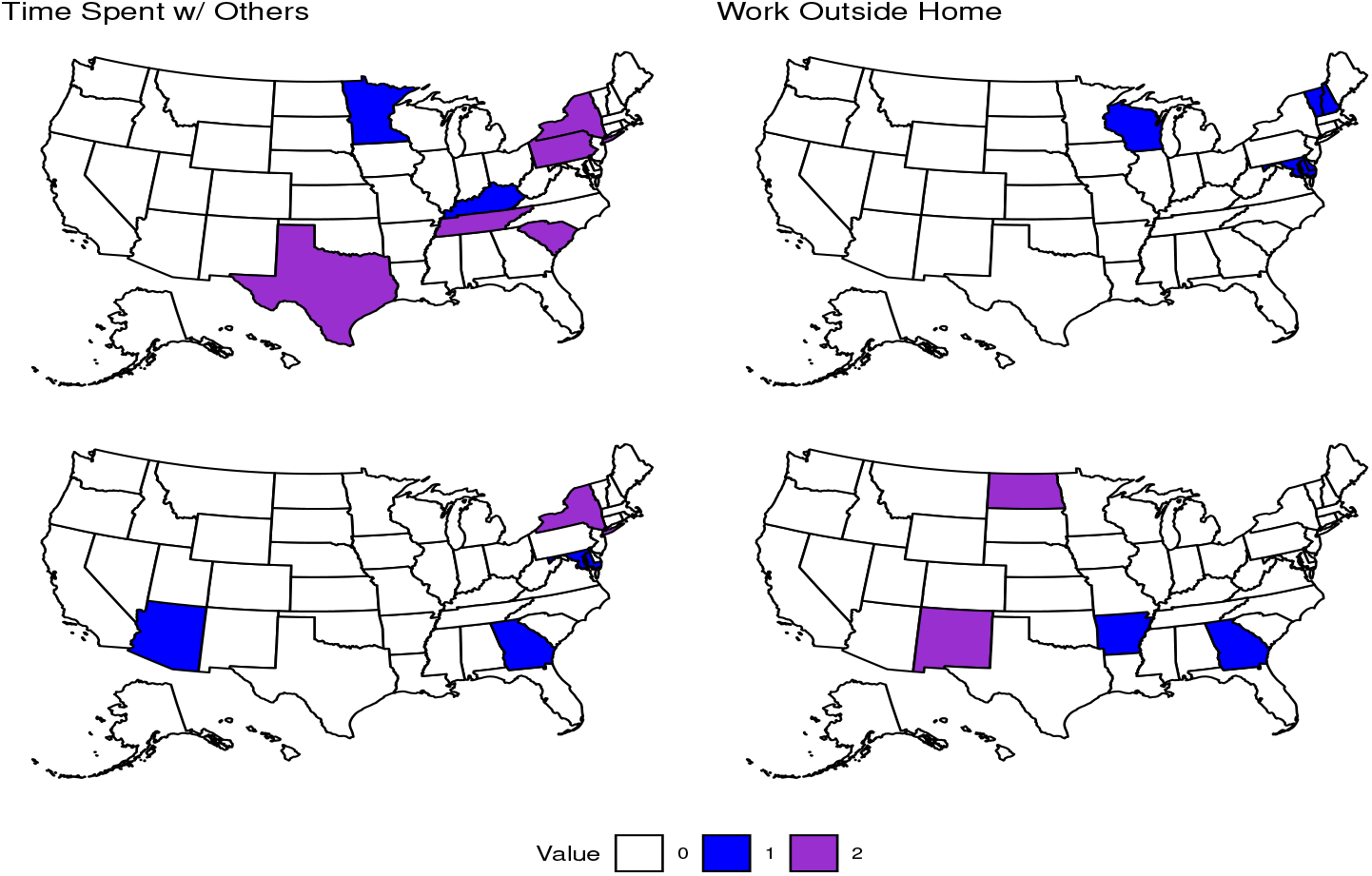
Effect of Social Isolation on Anxiety. States that had significantly positive or negative coefficients in their equation for anxiety for the first or second lag of variables related to Social Isolation in the first time period. The color depicts whether a state had a positive (blue), negative (purple), or no significant coefficient for the corresponding variable. TimeSpent lag 1: (positive) Kentucky, Minnesota, (negative) Pennsylvania, New York, South Carolina, Tennessee, Texas. Lag 2: (positive) Arizona, Georgia, Maryland, (negative) New York. Work lag 1: (positive) Delaware, Maryland, New Hampshire, Vermont, Wisconsin. Lag 2: (positive) Arkansas, Georgia, (negative) New Mexico, North Dakota.

There were five states that returned significant positive coefficients for the variable based on the estimated percentage of individuals that spent time with someone who they aren’t currently staying with in the past 24 hours and five states that returned significant negative coefficients for this variable. Our second social variable estimated the percentage of individuals that worked or went to school outside their home in the past 24 hours and gave significant positive results for seven states and significant negative results for two states. The initial results for the effect that our two social variables may have on anxiety indicate a varied, albeit limited response across the country.

**Figure 12:**
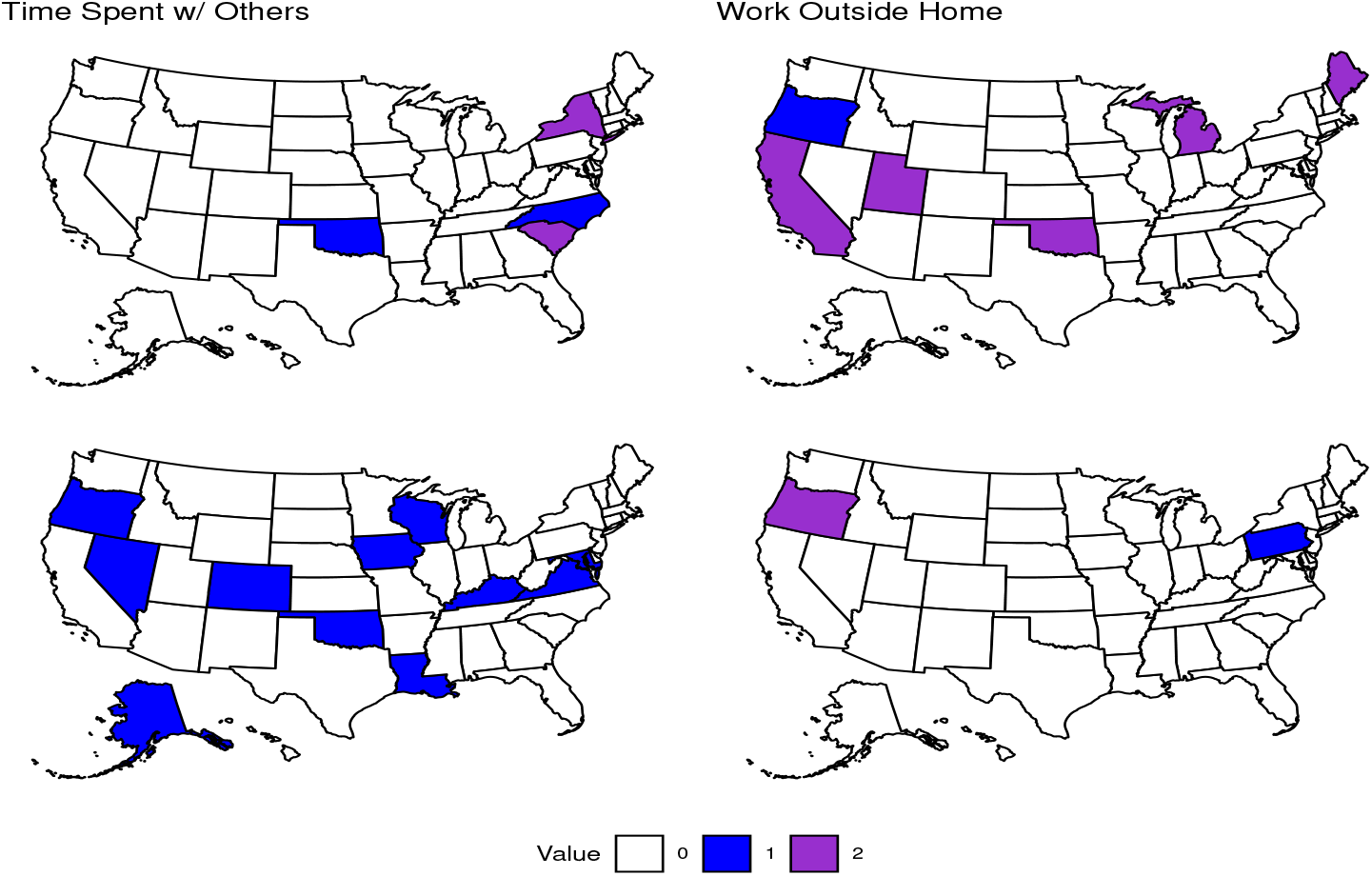
Effect of Social Isolation on Depression. States that had significantly positive or negative coefficients in their equation for depression for the first or second lag of variables related to Social Isolation in the first time period. The color depicts whether a state had a positive (blue), negative (purple), or no significant coefficient for the corresponding variable. TimeSpent lag 1: (positive) North Carolina, Oklahoma, (negative) New York, South Carolina. Lag 2: (positive) Alaska, Colorado, Iowa, Kentucky, Louisiana, Maryland, Nevada, Oklahoma, Oregon, Virginia, Wisconsin. Work lag 1: (positive) Oregon, (negative) California, Maine, Michigan, Oklahoma, Utah. Lag 2: (positive) Pennsylvania, (negative) Oregon.

Our chosen social variables seem to have more of an impact and a more consistent impact in equations for depression compared to anxiety in the first time period, with 12 states returning significant positive coefficients for the variable related to time spent with others and two states returning significant negative coefficients for this variable. Our second social variable related to individuals working outside their home yielded significant positive results for two states and significant negative results for six states. Note that one state had a significant positive coefficient in the first lag of this variable and a significant negative coefficient for the second lag. Overall, an increase in our first social variable generally led to an increase in anxiety across the country. In addition more states returned significant results in the second lag of this variable indicating that the impact of meeting new individuals may take days to present itself. Our second social variable had less of an impact and generally led to individuals becoming less depressed as more people started working outside their homes. Also, this variable seems to have a more immediate impact on depression as we see several more significant results in the first lag.

### B.3 Impulse Response Function Results

#### B.3.1 Effects of COVID-19 in the First Time Period

**Figure 13:**
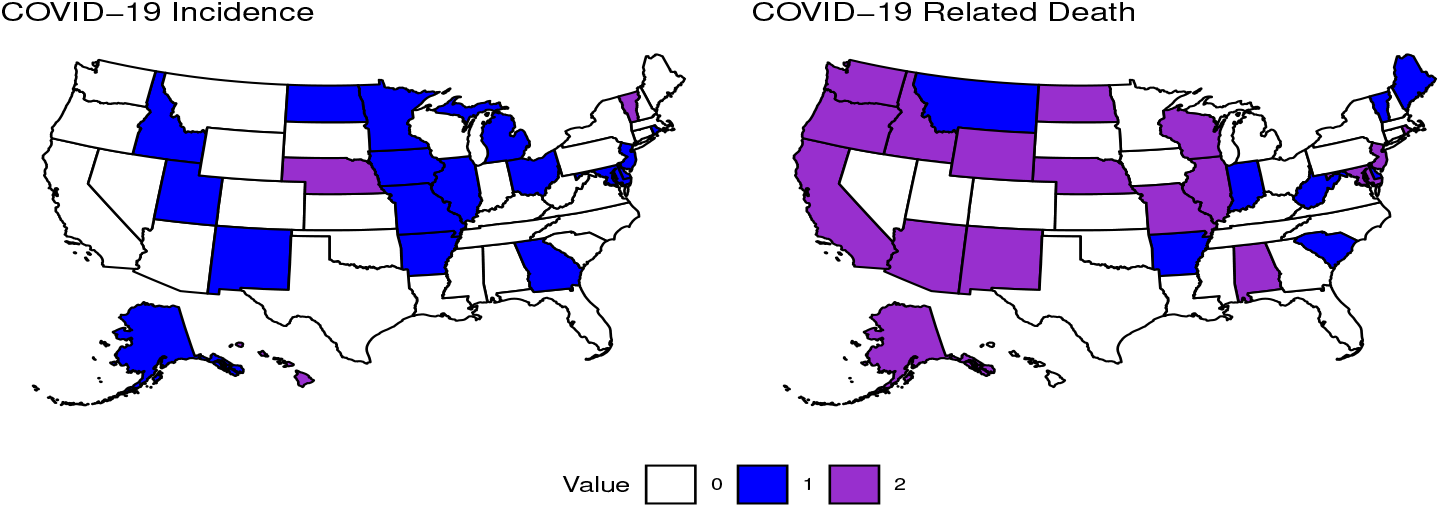
Effect of COVID-19 on Anxiety. States that had significantly positive or negative responses for variables related to COVID-19 in the first time period. The color depicts whether a state had a positive (blue), negative (purple), or no significant coefficient for the corresponding variable. COVID-19 Incidence: (positive) Alaska, Arizona, Georgia, Idaho, Illinois, Iowa, Minnesota, New Jersey, New Mexico, Rhode Island, (negative) NA. COVID-19 death: (positive) Arkansas, Indiana, West Virginia, (negative) Missouri, Nebraska, New Jersey, New Mexico.

Moving on to impulse response functions (IRFs), there were ten states that had significant positive responses in anxiety to an impulse in COVID-19 incidence. For COVID-19 related death, three states had significant positive responses and four states had significant negative responses. This may indicate that individuals are more anxious about becoming infected than dying to COVID-19.

**Figure 14:**
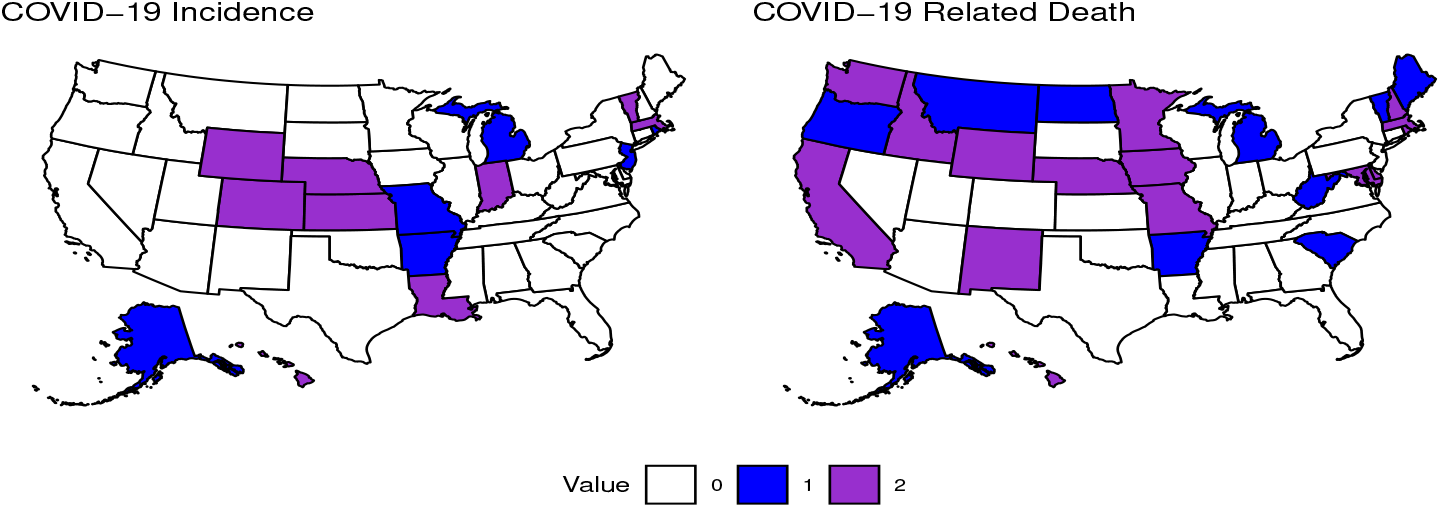
Effect of COVID-19 on Depression. States that had significantly positive or negative responses for variables related to COVID-19 in the first time period. The color depicts whether a state had a positive (blue), negative (purple), or no significant coefficient for the corresponding variable. COVID-19 Incidence: (positive) North Dakota, Rhode Island, (negative) Kansas, Kentucky, Vermont. COVID-19 death: (positive) Oregon, South Carolina, Vermont, West Virginia, (negative) Nebraska, Wyoming.

Two states had significant positive responses and 3 states had significant negative responses in depression to an impulse in COVID-19 incidence. Four states had significant positive responses and two states had significant negative responses in depression to an impulse in COVID-19 related death. This may indicate that either there is not a strong enough effect of these two variables on depression in the general population, or we have not effectively captured the signal with the current model.

#### B.3.2 Effects of Social Isolation in the First Time Period

**Figure 15:**
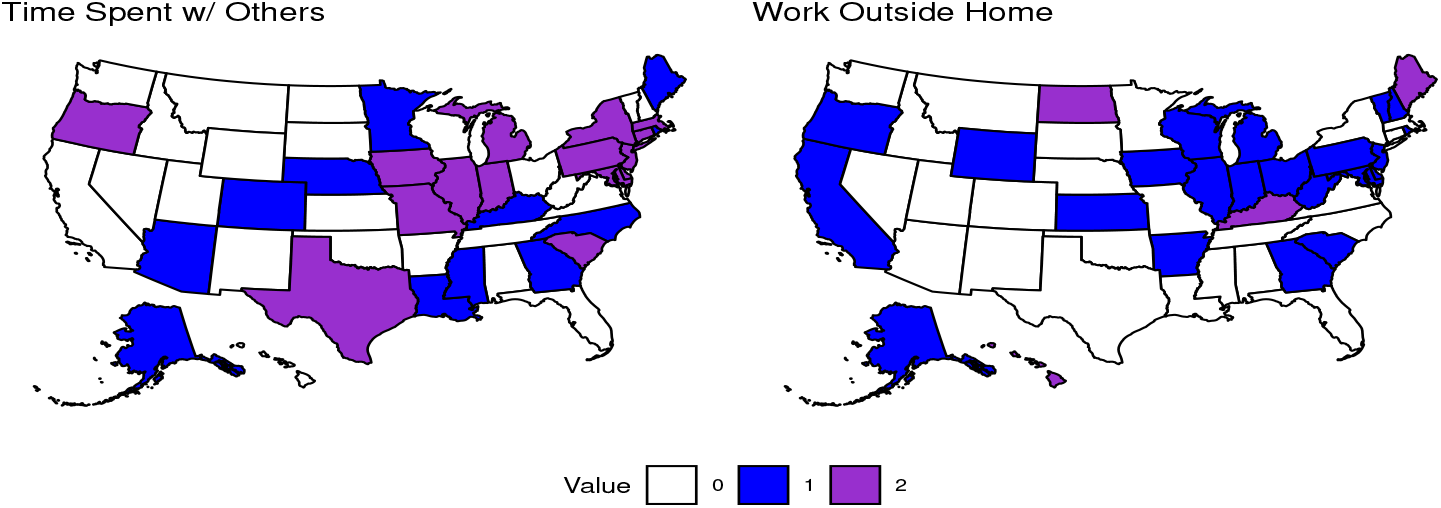
Effect of Social Isolation on Anxiety. States that had significantly positive or negative responses for variables related to Social Isolation in the first time period. The color depicts whether a state had a positive (blue), negative (purple), or no significant coefficient for the corresponding variable. TimeSpent: (positive) Alaska, Arizona, Colorado, Georgia, Louisiana, Nebraska, Maine, Minnesota, Mississippi, North Carolina, Rhode Island, Tennessee, (negative) Connecticut, Delaware, Illinois, Indiana, Iowa, Maryland, Massachusetts, Michigan, Missouri, New Jersey, New York, Oregon, Pennsylvania, South Carolina, Texas. Work: (positive) Alaska, Arkansas, California, Delaware, Georgia, Illinois, Indiana, Iowa, Kansas, Maryland, Michigan, New Hampshire, New Jersey, Ohio, Oregon, Pennsylvania, Rhode Island, South Carolina, Vermont, West Virginia, Wisconsin, Wyoming, (negative) Hawaii, Kentucky, Maine, North Dakota.

There were 12 states with significant positive responses and 15 states with significant negative responses in anxiety to an impulse in the time spent variable. An impulse in working outside your home led to 22 states with positive responses and four states with negative responses in anxiety. Overall, we see that more time spent with others can lead to a decrease in anxiety while working outside your home can lead to an increase.

**Figure 16:**
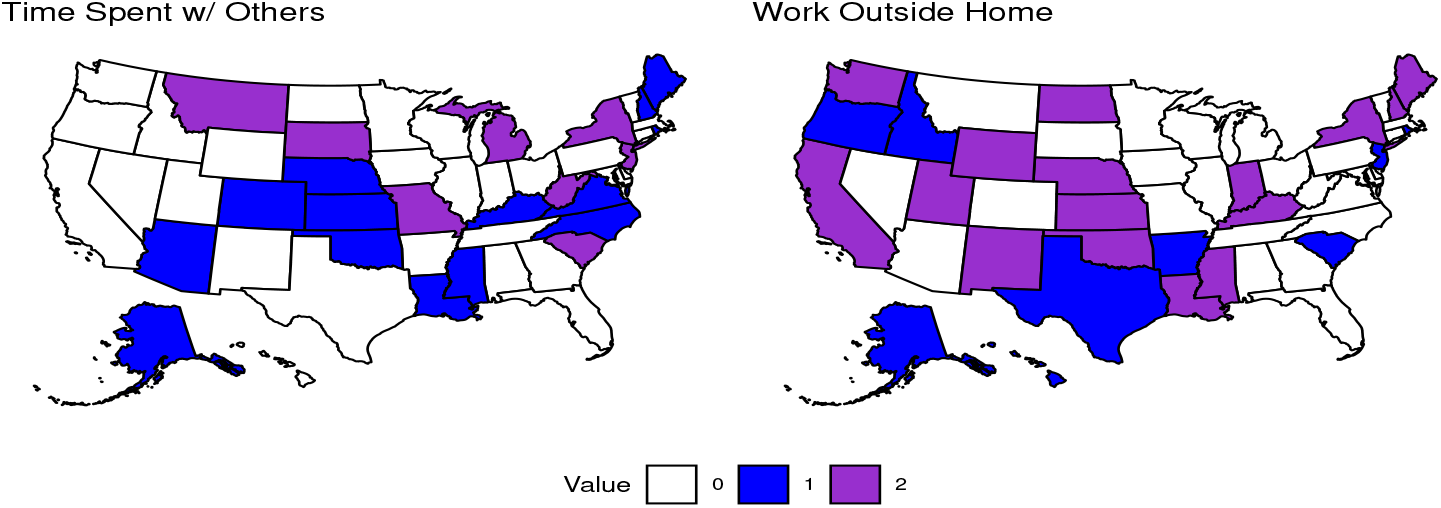
Effect of Social Isolation on Depression. States that had significantly positive or negative responses for variables related to Social Isolation in the first time period. The color depicts whether a state had a positive (blue), negative (purple), or no significant coefficient for the corresponding variable. Time-Spent: (positive) Alaska, Arizona, Colorado, Kansas, Kentucky, Louisiana, Maine, Mississippi, Nebraska, North Carolina, Oklahoma, Rhode Island, Vermont, Virginia, (negative) Michigan, Missouri, Montana, New Jersey, New York, South Carolina, South Dakota, West Virginia. Work: (positive) Alaska, Arkansas, Hawaii, Idaho, New Jersey, Oregon, Rhode Island, South Carolina, Texas, (negative) California, Indiana, Kansas, Kentucky, Louisiana, Maine, Mississippi, Nebraska, New Hampshire, New Mexico, New York, North Dakota, Oklahoma, Utah, Washington, Wyoming.

There are more significant results with these social variables compared to the variables related to COVID-19 for depression. Fourteen states yielded significant positive responses and 8 states yielded significant negative responses for depression with an impulse in the time spent variable. Nine states yielded significant positive responses and 16 states yielded significant negative responses for depression with an impulse in working outside the home. We see an opposite trend to what was observed with anxiety as there tends to be an increase in depression with an increase in the time spent variable. There is not as clear of a relationship between depression and working outside the home.

## C Effects of Vaccination in the Second Time Period

**Figure 17:**
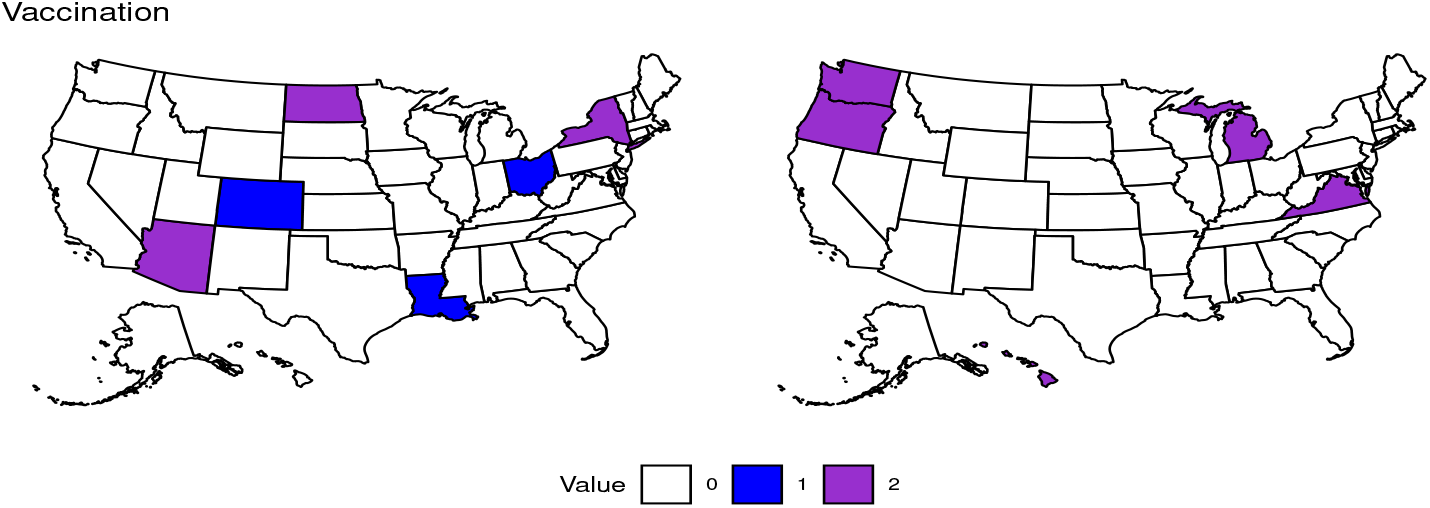
Effect of Vaccination on Anxiety. States that had significantly positive or negative coefficients in their equation for anxiety for the first or second lag of the variable for vaccination in the second time period. The color depicts whether a state had a positive (blue), negative (purple), or no significant coefficient for the corresponding variable. Vaccination Lag 1: (positive) Colorado, Louisiana, Ohio (negative) Arizona, New York, North Dakota. Vaccination Lag 2: (positive) NA, (negative) Hawaii, Michigan, Oregon, Virginia, Washington.

A total of three states returned significant positive coefficients for the first lag of vaccination indicating that increased levels of vaccination may have been associated with increased anxiety in those states during this period. A total of three states returned significant negative coefficients for the first lag of vaccination indicating that as vaccination increased, anxiety tended to decrease. No states returned significant positive coefficients for the second lag of vaccination. A total of five states returned significant negative coefficients for the second lag of vaccination in their respective equations for anxiety.

**Figure 18:**
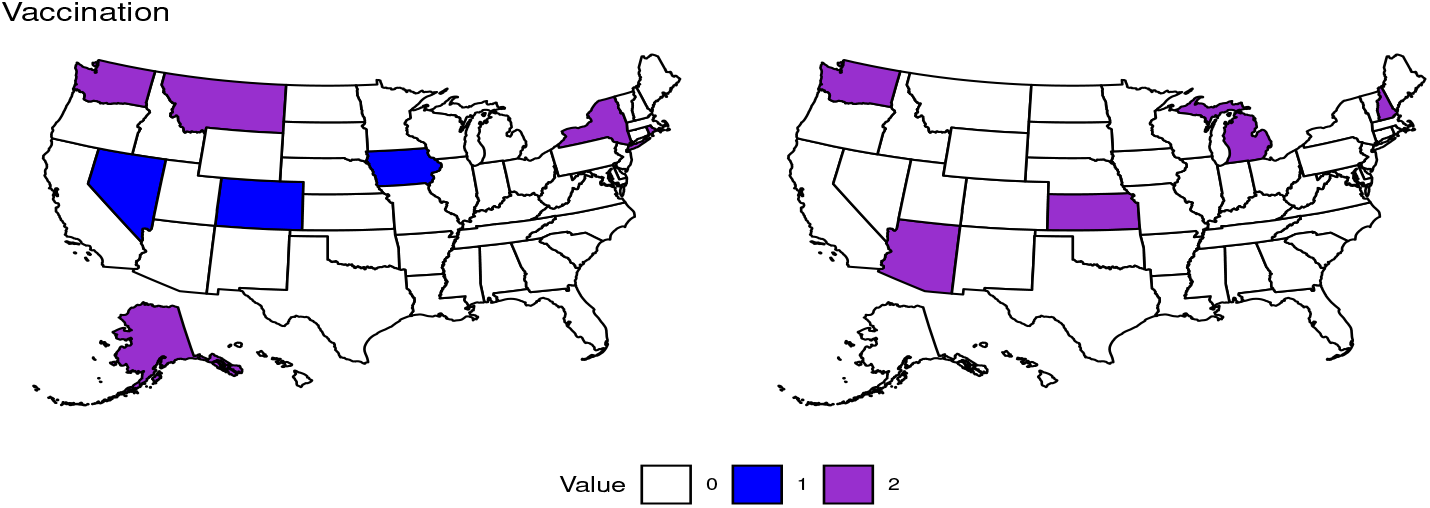
Effect of Vaccination on Depression. States that had significantly positive or negative coefficients in their equation for depression for the first or second lag of the variable for vaccination in the second time period. The color depicts whether a state had a positive (blue), negative (purple), or no significant coefficient for the corresponding variable. Vaccination Lag 1: (positive) Colorado, Iowa, Nevada (negative) Alaska, Montana, New York, Rhode Island, Washington. Vaccination Lag 2: (positive) NA, (negative) Arizona, Kansas, Michigan, New Hampshire, Washington.

Results show that three states returned significant positive coefficients for vaccination in their respective equations for depression, while five states returned significant negative coefficients for this variable. No states returned significant positive coefficients for vaccination, similar to the case with anxiety. On the other hand, five states returned significant negative coefficients for vaccination in their equations for depression.

**Figure 19:**
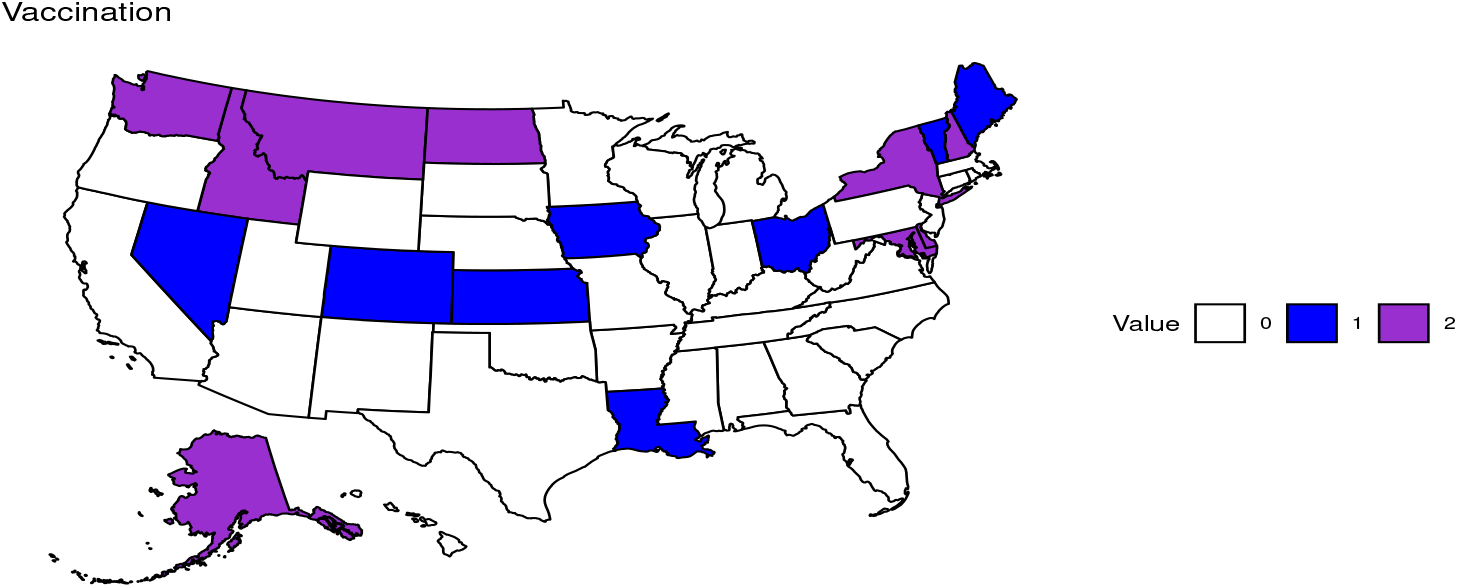
Effect of Vaccination on Anxiety. States that had significantly positive or negative responses for variables related to vaccination in the second time period. The color depicts whether a state had a positive (blue), negative (purple), or no significant coefficient for the corresponding variable. Vaccination: (positive) Colorado, Kansas, Louisiana, Maine, Ohio, Vermont, (negative) Arizona, Maryland, New York, North Dakota.

An impulse in vaccination had a varied effect across the country with six states having significant positive responses and four states with significant negative responses in anxiety.

**Figure 20:**
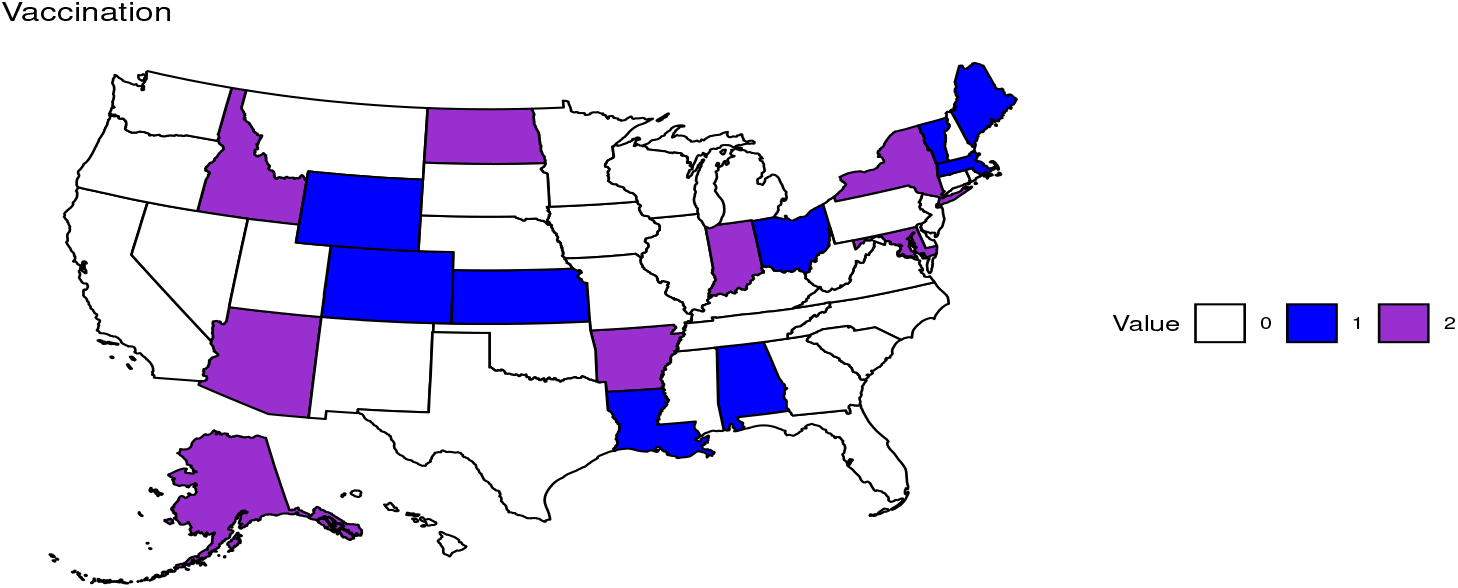
Effect of Vaccination on Depression. States that had significantly positive or negative responses for variables related to vaccination in the second time period. The color depicts whether a state had a positive (blue), negative (purple), or no significant coefficient for the corresponding variable. Vaccination: (positive) Alabama, Colorado, Kansas, Louisiana, Maine, Massachusetts, Ohio, Vermont, Wyoming, (negative) Alaska, Arizona, Arkansas, Idaho, Indiana, Maryland, New York, North Dakota.

Nine states had positive responses and 8 states had negative responses for depression to an impulse in vaccination.

## References

[1] World Health Organization (WHO), “COVID-19 Dashboard.” Geneva: World Health Organization, 2020. Available online: https://covid19.who.int/ (Accessed: 03-06-2022).

[2] Centers for Disease Control and Prevention (CDC), “COVID-19 Overview and Infection Prevention and Control Priorities in non-U.S. Healthcare settings.” Available online: https://www.cdc.gov/coronavirus/2019-ncov/hcp/non-us-settings/overview/index.html\# (Accessed: 03-06-2022).

[3] Centers for Disease Control and Prevention (CDC)“Mental Health, Substance Use, and Suicidal Ideation During the COVID-19 Pandemic — United States, June 24–30, 2020” Available online: https://www.cdc.gov/mmwr/volumes/69/\wr/pdfs/mm6932a1-H.pdf (Accessed: 03-06-2022).

[4] World Health Organization (WHO), “Coronavirus disease (COVID-19): How is it transmitted?” Geneva: World Health Organization, 2020. Available online: https://www.who.int/news-room/questions-and-answers/item/coronavirus-disease-covid-19-how-is-it-transmitted (Accessed: 03-22-2022).

[5] J. L. St. Sauver, G. S. Lopes, W. A. Rocca, K. Prasad, M. R. Majerus, A. H. Limper, D. J. Jacobson, C. Fan, R. M. Jacobson, L. J. Rutten, A. D. Norman, C. M. Vachon, “Factors Associated With Severe COVID-19 Infection Among Persons of Different Ages Living in a Defined Midwestern US Population.” Mayo Clinic Proceedings, vol. 96, pp. 2528–2539, issue 10, 2021.

[6] P. Nouvellet, S. Bhatia, A. Cori, K. E. C. Ainslie, et al. “Reduction in mobility and COVID-19 Transmission.” Nat Commun, vol. 12, pp. 1090, 2021.

[7] M. V. Fasano, M. Padula, M. Azrak, A. J. Avico, M. Sala, M. F. Andreoli, “Consequences of Lockdown During COVID-19 Pandemic in Lifestyle and Emotional State of Children in Argentina.” Front. Pediatr., 2021. Available online: 10.3389/fped.2021.660033 (Accessed: 03-08-2022).

[8] D. Richter, S. Riedel-Heller, S. Zürcher, “Mental health problems in the general population during and after the first lockdown phase due to the SARS-Cov-2 pandemic: Rapid review of multi-wave studies.” Epidemiology and Psychiatric Sciences, vol. 30, p. e27, 2021.

[9] X. Nie, K. Feng, S. Wang, Y. Li, “Factors Influencing Public Panic During the COVID-19 Pandemic.” Front Psychol., vol. 12, p. e576301, 2021.

[10] J. S. Novotný, J. P. Gonzalez-Rivas, Š Kunzová, M. Skladaná, A. Pospíšilová, A. Polcrová, J. R. Medina-Inojosa, F. Lopez-Jimenez, Y. E. Geda, G. B. Stokin, “Risk Factors Underlying COVID-19 Lockdown-Induced Mental Distress.” Front. Psychiatry, vol. 11, p. e603014, 2020.

[11] M. É. Czeisler, R. I. Lane, E. Petrosky, et al. “Mental Health, Substance Use, and Suicidal Ideation During the COVID-19 Pandemic — United States, June 24-30, 2020.” MMWR Mor Mortal Wkly Rep, vol. 69, pp. 1049–1057, 2020.

[12] A. Novotney, “The risks of social isolation.” Monitor on Psychology, vol. 50, no. 5, 2019.

[13] A. Pearman, M. L. Hughes, E. L. Smith, S. D. Neupert, “Mental Health Challenges of United States Healthcare Professionals During COVID-19.” Front. Psychol., vol. 11, p. e2065, 2020.

[14] M. M. Trivedi, A. Das, “Did the Timing of State Mandated Lockdown Affect the Spread of COVID-19 Infection? A County-level Ecological Study in the United States.” J Prev Med Public Health, vol. 54, no. 4, pp. 238–244, 2021.

[15] W. J. Hennigan, A. Park, J. Ducharme, “The U.S. Fumble Its Early Vaccine Rollout. Will the Biden Administration Put America Back on Track?” Time, 2021. Available online: https://time.com/5932028/vaccine-rollout-joe-biden/ (Accessed: 04-10-2022).

[16] C. Wilson, “The U.S. COVID-19 Vaccine Rollout is Getting Faster. But is it Fast Enough?” Time, 2021. Available online: https://time.com/5938128/covid-19-vaccine-rollout-biden/ (Accessed: 04-10-2022).

[17] M. C. Staff, “Herd immunity and COVID-19: What you need to know.” Mayo Clinic, 2021. Available online: https://www.mayoclinic.org/diseases-conditions/coronavirus/in-depth/herd-immunity-and-coronavirus/art-20486808 (Accessed: 04-10-2022).

[18] A. Reinhart, L. Brooks, M. Jahja, et al. “An open repository of real-time COVID-19 indicators.” Proceedings of the National Academy of Sciences, vol. 118, p. e2111452118, 2021.

[19] H. Lütkepohl, “A New Introduction to Multiple Time Series Analysis.” Springer, 2005.

[20] J. Stock, and M. Watson. “Vector Autoregressions.” Journal of Economic Perspectives. vol. 15(4), pp. 101–115, 2001.

[21] B. Pfaff, “Analysis of Integrated and Cointegrated Time Series with R. Second Edition.” Springer, New York, 2008. ISBN 0-387-27960-1

[22] B. Pfaff, “VAR, SVAR and SVEC Models: Implementation Within R Package vars.” Journal of Statistical Software, vol. 27, no. 4, 2008.

[23] T. Hwang, K. Rabheru, C. Peisah, W. Reichman, M. Ikeda, “Loneliness and social isolation during the COVID-19 pandemic.” International Psychogeriatrics. vol. 32(10), pp. 1217–1220, 2020.

[24] L. Ramiz, B. Contrand, M. Y. R. Castro, M. Dupuy, L. Lu, C. Sztal-Kutas, E. Lagarde, “A longitudinal study of mental health before and during COVID-19 lockdown in the French population.” Globalization and Health, vol. 17, pp. 29, 2021.

[25] I. Magnúsdóttir, A. Lovik, A. B. Unnarsdóttir, D. McCartney, et al. “Acute COVID-19 severity and mental health morbidity trajectories in patient populations of six nations: an observational study.” The Lancet Public Health, 2022.

[26] H. Mautong, J. Gallardo-Rumbea, G. Alvarado-Villa, J. Fernández-Cadena, D. Andrade-Molina, C. Orellana-Román, I. Cherrez-Ojeda, “Assessment of depression, anxiety and stress levels in the Ecuadorian general population during social isolation due to the COVID-19 outbreak: a cross-sectional study.” BMC Psychiatry, vol. 21, pp. 212, 2021.

[27] N. Vindegaard, M. E. Benros, “COVID-19 pandemic and mental health consequences: Systematic review of the current evidence.” Brain, Behavior, and Immunity, vol. 89, pp. 531–542, 2020.

[28] J. Xiong, O. Lipsitz, F. Nasri, L. M. W. Lui, et al. “Impact of COVID-19 pandemic on mental health in the general population: A systematic review.” J Affect Disord, vol. 277, pp. 55–64, 2020.

[29] F. Kämpfen, I. Kohler, A. Ciancio, W. Bruine de Bruin, J. Maurer, H. -P. Kohler, “Predictors of mental health during the Covid-19 pandemic in the US: Role of economic concerns, health worries and social distancing.” PLoS One, vol. 15, p. e0241895, 2020.

[30] T. Wu, X. Jia, H. Shi, J. Niu, X. Yin, J. Xie, X. Wang, “Prevalence of mental health problems during the COVID-19 pandemic: A systematic review and metaanalysis.” Journal of Affective Disorders, vol. 281, pp. 91–98, 2021.

[31] Z. Santini, P. E. Jose, E. Y. Cornwell, A. Koyanagi, L. Nielsen, et al. “Social disconnectedness, perceived isolation, and symptoms of depression and anxiety among older Americans (NSHAP): a longitudinal mediation analysis.” The Lancet Public Health, vol. 5, pp. e62–e70, 2020.

[32] B. Saddik, A. Hussein, A. Albanna, I. Elbarazi, A. Al-Shujairi, et al. “The psychological impact of the COVID-19 pandemic on adults and children in the United Arab Emirates: a nationwide cross-sectional study.” BMC Psychiatry, vol. 21, pp. 224, 2021.

[33] D. Lekkas, J. A. Gyorda, G. D. Price, Z. Wortzman, N. C. Jacobson, “Using the COVID-19 Pandemic to Assess the Influence of News Affect on Online Mental Health-Related Search Behavior Across the United States: Integrated Sentiment Analysis and the Circumplex Model of Affect.” Journal of Medical Internet Research, vol. 24, pp. e32731, 2022.

[34] A. Burton, A. McKinlay, H. Aughterson, D. Fancourt, “Impact of the COVID-19 pandemic on the mental health and well-being of adults with mental health conditions in the UK: a qualitative interview study.” Journal of Mental Health, pp.1-8, 2021.

[35] Z. Su, D. McDonnell, J. Wen, M. Kozak, J. Abbas, et al. “Mental health conse-quences of COVID-19 media coverage: the need for effective crisis communication practices.” Global Health, vol. 17, 2021.

[36] A. Olagoke, O. Olagoke, A. Hughes, “Exposure to coronavirus news on mainstream media: The role of risk perceptions and depression.” British Journal of Health Psychology. vol. 25(4), pp. 865–874, 2020.

[37] A. Adeel, M. Catalano, O. Catalano, et al. “COVID-19 Policy Response and the Rise of the Sub-National Governments” Canadian Public Policy. vol. 46(4), pp. 565–584, 2020.

